# Minimally processed cow’s milk and early childhood asthma-related outcomes: exploratory findings from the early terminated MARTHA randomized trial

**DOI:** 10.64898/2026.10.23.26363903

**Authors:** Melanie Weber, Yvi Huynh, Giulia Pagani, Martin Depner, Silvia Gschwendtner, Hans Demmelmair, Chrysanthi Skevaki, Hermelijn H. Smits, Kasper Hettinga, Michael Schloter, Christine Strunz-Lehner, Joachim Heinrich, Erika von Mutius, Markus J. Ege

## Abstract

**Background:** Raw milk consumption has repeatedly been shown to be inversely related to asthma, respiratory infections, and allergies but carries a potential risk of life-threatening infections. The MARTHA trial (DRKS00014781) used minimally-processed pasteurized milk as a microbiologically safe proxy for raw milk and compared it against ultra-heat-treated milk for efficacy on atopic sensitization, asthma, and respiratory infections.

**Methods:** Healthy children were recruited through birth clinics in Germany. At 6 to 12 months, they were centrally randomized to obtain minimally-processed full-cream versus ultra-heat-treated semi-skimmed cow’s milk daily until age 3 years. During the SARS-CoV-2 pandemic, recruitment was stopped, and the study was continued without confirmatory intention. The primary endpoint was shifted from asthma as composite parent-reported diagnosis of asthma or recurrent obstructive bronchitis until age 5 to atopic sensitization at age 3 years. Participants, caregivers, and study staff were blinded to group assignments.

**Results:** We randomized 130 individuals into each study arm. The intervention was unrelated to atopic sensitization (risk ratio, RR=1.13 [0.67-1.89]), which occurred in 28 individuals of the intervention (n=99) and 24 of the comparator group (n=96). Asthma showed a trend for an inverse association (hazards ratio, HR=0.55 [0.27−1.16], n=252), which was pronounced in the 82 children with a family history of asthma (post-hoc HR=0.30 [0.09−0.93]). The intervention was inversely associated with rhinitis and cough in the older half of the children (post-hoc RR=0.82 [0.67-0.998] and RR=0.75 [0.60-0.94], respectively). No adverse events were judged intervention-related.

**Interpretation:** The intervention did not reduce atopic sensitization at age 3 years. Exploratory analyses suggested possible protective effects on asthma and respiratory symptoms, particularly in children with parental asthma and in those less affected by pandemic-related disruptions. These findings justify but do not replace a definitive trial.

**Funding:** Dutch Longfonds; Bavarian State-Ministry of Health, Care and Prevention; German Center for Lung Research (DZL); FrieslandCampina.

## Introduction

Children from rural areas growing up on farms are at a significantly lower risk for developing asthma, allergic rhinoconjunctivitis and allergic sensitization to inhalant and food allergens than their classmates. ^1,2^ This relationship has been shown in many populations and is sustained into adult life.^1,2^ The farm exposures contributing to the reduced risk of asthma and allergies have been identified as contact with livestock, contact with animal feed, and the consumption of unprocessed cow’s milk.^1,3–10^

The inverse association of asthma, allergies and consumption of unprocessed milk directly from a farm has been replicated in various independent studies across many countries and was found independently of other potentially confounding exposures.^11^ A meta-analysis including comparable exposure and outcome definitions, i.e. consumption of farm milk during the first year of life and childhood-onset asthma, showed consistent inverse associations.^11^

The aim of this study was to prove the protective effect of *minimally-processed full-cream cow’s milk* compared to *ultra-heat treated semi-skimmed milk* on childhood asthma, allergies, respiratory infections, and low-grade inflammation.

Minimally-processed milk has a normal fat content of about 4%, is not centrifuged for separating the fat fraction, and not homogenized. This minimally-processed product has been pasteurized according to legal requirements for pasteurized milk.

The comparator consisted of semi-skimmed, ultra-heat-treated (UHT) milk. Mostly UHT milk is sold with a reduced fat content of 1.5%. Semi-skimmed UHT milk can be stored at room temperature and is widely used, particularly in Germany, where we found a proportion of 62% among infants who were fed with cow’s milk in the first year of life.^12^ Semi-skimmed UHT milk was also the reference milk type in many of the above-mentioned farm studies and thus used for sample size calculation.

The primary outcome was atopic sensitization at age 3 years, defined as specific IgE ≥ 0.7kU/l to food or inhalant allergens. The participants were healthy children of both sexes enrolled at age 6 to 12 months.

## Methods

### Trial design

This trial investigated superiority of minimally processed, pasteurized, full-cream cow’s milk over UHT treated semi-skimmed milk for the prevention of asthma, atopy, and infections. The design was a randomized, double-blind, placebo-controlled, two-arm parallel-group prevention trial with a nutritional intervention. The allocation ratio was 1:1.

During the SARS-CoV-2 pandemic, the following important changes to methods after trial initiation occurred: The treatment regimen was changed on 2020-09-29 from powdered milk products to liquid milk because the provider stopped delivery. Both intervention and comparator milk were shipped on cool packs for blinding. Due to delayed recruitment, the primary outcome was changed on 2021-06-01 from asthma to atopic sensitization because the latter is more frequent and can be measured at 3 years. The sample size calculation was adjusted to atopic sensitization, which resulted in a target sample size of 956 individuals instead of the 2700 individuals needed for asthma. Because of shortage of staff during the first SARS-CoV-2 wave, the study center Regensburg was closed in April 2020 and not reopened. The 9 individuals so far included were cared for by the single remaining center Munich.

Because funding was discontinued, recruitment was stopped on 2021-10-04 and the trial was continued with 260 randomized individuals without confirmatory intention upon recommendation by the data safety monitoring board (DSMB). Consequently, calculation of sample size was not revised.

The study and all changes were approved by the ethics committee at the Medical Faculty of LMU Munich (18-0405) and registered with the German clinical trials registry (DRKS) under the trial number DRKS00014781 (universal trial number U1111–1216-7580).

### Participants

Subjects must meet all of the following inclusion criteria to be eligible for enrolment: healthy infants of both sexes born after at least 35 weeks of gestation; age 6 to 12 months at randomization; caregivers with the ability to follow study instructions and likely to attend and complete all required visits and the web-based diary; written informed consent of the parents or guardians.

Subjects presenting with any of the following exclusion criteria were not included in the trial: suspected or confirmed cow’s milk allergy, milk intolerance, lactose intolerance, suspected immunodeficiency, cystic fibrosis, failure to thrive; simultaneous participation in another clinical trial with a conflicting intervention; a sibling already participating in this trial; maternal age below 18 years; uncertainty of the investigator about prospective adherence to study protocol, e.g. known or persistent abuse of medication, drugs or alcohol in caregivers.

The study was carried out in the two study centers Munich and, until April 2020, in Regensburg, Germany. Families-to-be were approached in academic and non-academic birth clinics in the Munich metropolitan area and in Regensburg. Clinical visits were performed at a central outpatient clinic in Munich and Regensburg. During shutdown, few clinical visits were performed as video conferences without biosampling. The specific settings of the SARS-CoV-2 pandemic with the corresponding lockdown measures could have influenced the observed results by rare respiratory infections and rebound effects.

### Interventions

The intervention consisted of a daily dose of pasteurized, non-homogenized, non-separated, full-cream cow’s milk with a natural fat content of 4.0% in the intervention arm and ultra-heat-treated, homogenized milk with a fat content of 1.5% in the comparator arm.

The intervention started after weaning and not before 6 months of age and lasted until age 36 months or the third clinical visit whatever was first. Children between 6 to 9 months consumed 200 mL / day; children 10 months and older consumed 2 x 140 mL portions. The daily milk serving had to be consumed by the study child within 1 hour. The milk was consumed purely or as a cereal meal without any heating. As a marker for adherence, we assessed the fatty acid composition in erythrocyte membranes during the intervention.^13,14^

### Outcomes

The primary outcome atopic sensitization at 3 years was defined as specific IgE ≥ 0.7 kU/l to any food or inhalant allergen as measured in the Central Laboratory of LMU Hospitals using their standard method.^15^ Asthma was defined as onset of parent-reported diagnosis of asthma or recurrent bronchitis until age 5 years according to the German version of the ISAAC definition.^16^ Atopic eczema was assessed as onset of parent-reported atopic eczema until age 3 years. Low-grade inflammation at age 3 years was measured by serum levels of high-sensitivity C-reactive protein (hs-CRP).^17^ Respiratory tract infections were measured by the weekly presence of rhinitis and cough until age 3 years. Other infection-related outcomes such as otitis media were not assessed because they were extremely rare due to lack of exposure.

With the above-described necessary adjustments for sample size and study duration, the original secondary outcome of atopic sensitization at 3 years became the primary outcome. In children who had to terminate the intervention before age 3 years because of the termination of the entire trial, the final visit was preponed by up to 10 months.

The original primary outcome asthma until age 5 years was used as a secondary outcome. Since the last clinical visit had to occur at 3 years at the latest, asthma was not clinically assessed but replaced by the well-established ISAAC definition.^16^ To adequately consider drop-outs, it was operationalized as time-to-onset of a parent-reported physician’s diagnosis of asthma or parent-reported recurrent bronchitis with episodes in at least two different quarterly questionnaires. Likewise, atopic eczema was operationalized as time-to-onset of a parent-reported physician’s diagnosis of atopic eczema.

Adverse events have been reported previously.^15^ An adverse event (AE) was defined as any gastrointestinal, respiratory, or cutaneous symptoms compatible with milk allergy or intolerance. A serious adverse event (SAE) was defined as any condition resulting in death, life-threatening situations, unscheduled hospitalizations of more than 3 days, hospitalizations due to COVID-19, or failure to thrive.^15^ Information about symptoms was collected by weekly diaries and evaluated by study physicians by phone calls or, if necessary, additional clinical visits with sIgE measurement, skin prick tests, and food provocation tests under clinical conditions.^15^ The clinical team including principal investigator and study physicians decided whether an adverse event was related to the intervention. All assessors were blinded to the intervention group.

### Sample size

The expected effect size was estimated by the effect of raw versus shop milk consumed during 1st year of life on atopic sensitization based on data from the PARSIFAL study (n=12,070, OR=0.67 [0.47–0.96]) and the GABRIELA study (n=6,191, OR=0.68 [0.57-0.82]).^9,10^

A single stage (fixed sample size) design was chosen. Based on cohort data (Robert Koch Institute 2019) and the literature review (Brick 2020), we assumed for the primary outcome (atopy at age of three years) a prevalence of 25% in the control and of 17% in the intervention group (Relative Risk 0.68). A total of 406 subjects per group were needed to detect this difference on a two-sided significance level of 5% with a power of 80%. To account for 15% drop-outs we added additional subjects. This resulted in (2x406)/0.85 ≈ 956 subjects who had to be recruited into the trial.

### Randomization

Upon request by the site staff members, randomization was performed through the online data collection system “CASTOR”, which was certified for clinical studies and approved by the data protection officer of LMU Hospitals. The randomization was based on a permuted balanced block design with random block length. Individuals were allocated to 8 different groups identified by milk identifiers, which were used together with address data for shipment of milk products. The 8 groups comprised 4 groups receiving the intervention milk and 4 groups receiving the comparator. Neither the investigators nor the family were informed to which study arm the infant was allocated. The procedure considered stratification by study site. Emergency unblinding of individual study participants was not considered.

The CASTOR system used different roles for study staff. While the clinical team did not have access to randomization data, the team dedicated to milk shipment could not access clinical data of the participants.

The random allocation sequence was generated automatically by the CASTOR tool. The clinical team (physicians and study nurses) enrolled participants. The milk shipment team downloaded the assignment to the study arm from the CASTOR system and kept an allocation list on a protected section of the server of LMU Hospitals.

### Blinding

The participating children and their caregivers, all members of the coordinating team and the clinical teams were blinded to group allocation. Blinding of the milk was achieved by using the same expiry date and cooled shipping of both milk types by the same distributor. Furthermore, information about the manufacturers was not provided to the participating families; packaging did not contain any recognizable reference of group allocation. The DSMB had the right to be unblinded if necessary but did not make use of it. The secondary outcomes asthma, atopic eczema, and infections were reported by the parents.

### Statistical methods

All data were double pseudonymized by an external data trustee before statistical analysis. All analyses were performed using R 4.5.1. Distribution of demographic variables and baseline variables was compared by standardized differences (SD, R package *tableone*), and SD<0.1 was considered acceptable. Because of the unequal distribution of baseline variables, inverse probability of treatment weighting (IPTW) was performed for average treatment effects using the R package *survey*. For this purpose, a propensity score (PS) was calculated as the predicted probability from a logistic regression for assignment to the intervention arm versus the comparator arm modeled by variables listed in Supplemental Figure 1. Missing data in the variables used for the PS were imputed by multiple imputation (R package mice, m=5). Stabilized weights were calculated as 0.5/PS for the intervention arm and 0.5/(1-PS) for the comparator arm. Balancing of IPTW was verified by SD<0.1. Descriptive analyses included a comparison of the study arms with respect to frequencies or mean values with standard deviation. Numbers of participants from weighted analyses are rounded to integer values. Birth dates and age calculation was rounded to weeks.

While item nonresponse did not occur due to forced completion of electronic questionnaires, module nonresponse arose from missed biosampling or questionnaires. Questionnaires collected during phases with a low frequency of circulating viruses were excluded for all participants (variable-wise deletion). These phases were defined by a weekly rhinitis prevalence below 30% and occurred during lockdown measures or holiday seasons. During holidays milk distribution was discontinued for a maximum of 3 weeks.

When participants missed 4 consecutive surveys, they were considered dropouts and censored at the last completed survey. Missing data for sIgE and hs-CRP measurements were treated by inverse probability of selection weighting (IPSW) for module nonresponse. The resulting weights were multiplied by the IPTW weights, and weighted association measures were reported. The percentage of complete data available was described per baseline variable.

The primary outcome of atopic sensitization was assessed by a risk ratio calculated by a survey-weighted generalized linear Poisson model.

The secondary outcomes of asthma and atopic eczema were assessed by time-to-event analyses in intention-to-treat, thereby considering dropouts as censored. Time-to-event analyses were performed using weighted Kaplan-Meier estimates and weighted Cox proportional hazards models based on quarterly data from the diaries.

The secondary outcome hs-CRP at 3 years was compared between study arms by a geometric means difference with confidence intervals in an intention-to-treat IPTW/IPSW analysis.

The secondary outcomes related to infections were assessed on a weekly basis. Risk ratios for rhinitis and cough were modeled by the intervention using survey-weighted generalized linear Poisson models adjusting for the design effect by multiple observations per individual.

Sensitivity analyses were used to explore cohort effects on infections as children entered lockdown and post-lockdown phases at different ages. A potential effect modification of the association of the intervention with respiratory infections by a cohort effect was explored by including an interaction term with birth date dichotomized at the median (2019-12-16).

Stratified analyses were prespecified for fatty acid blood levels before intervention and polymorphisms in the *FADS1/2* genes. Since the respective measurements could not be performed due to budget cuts, we used family history of asthma as a surrogate marker of genotypes for post-hoc subgroup analyses.

Ancillary analyses on the gut microbiome evaluated a butyrate-producer maturation score using the published PASTURE prediction model for butyrate producing bacteria (see online methods).^18^

## Results

Recruitment started 2019-07-29 and was terminated on 2021-10-04 due to slow recruitment and lack of funding. The decision was made by the principal investigators upon the suggestion of the DSMB. The funding agency did not play a role in the decision. The last clinical visit took place on 2023-06-26, and follow-up was continued by electronic questionnaires until age 5 years, which was reached by the youngest participant on 2026-02-22.

Of all 260 randomized individuals, 195 were assessed for the primary outcome and 252 were assessed for the secondary outcomes asthma and atopic eczema (Figure 1). Applying IPTW, demographic and clinical characteristics at baseline were well balanced between the study arms (Table 1, Supplemental Figure 1). In 82 participants (32%), at least one parent suffered from asthma. On average, children participated 109 weeks (about 2 years) in the intervention (Supplemental Table 1). Additional milk was consumed for at least 25 weeks in about half of the children (Supplemental Table 1). Potential harms were assessed by 22,988 surveys for all 260 randomized individuals for a mean of 109 weeks.

**Figure 1:**
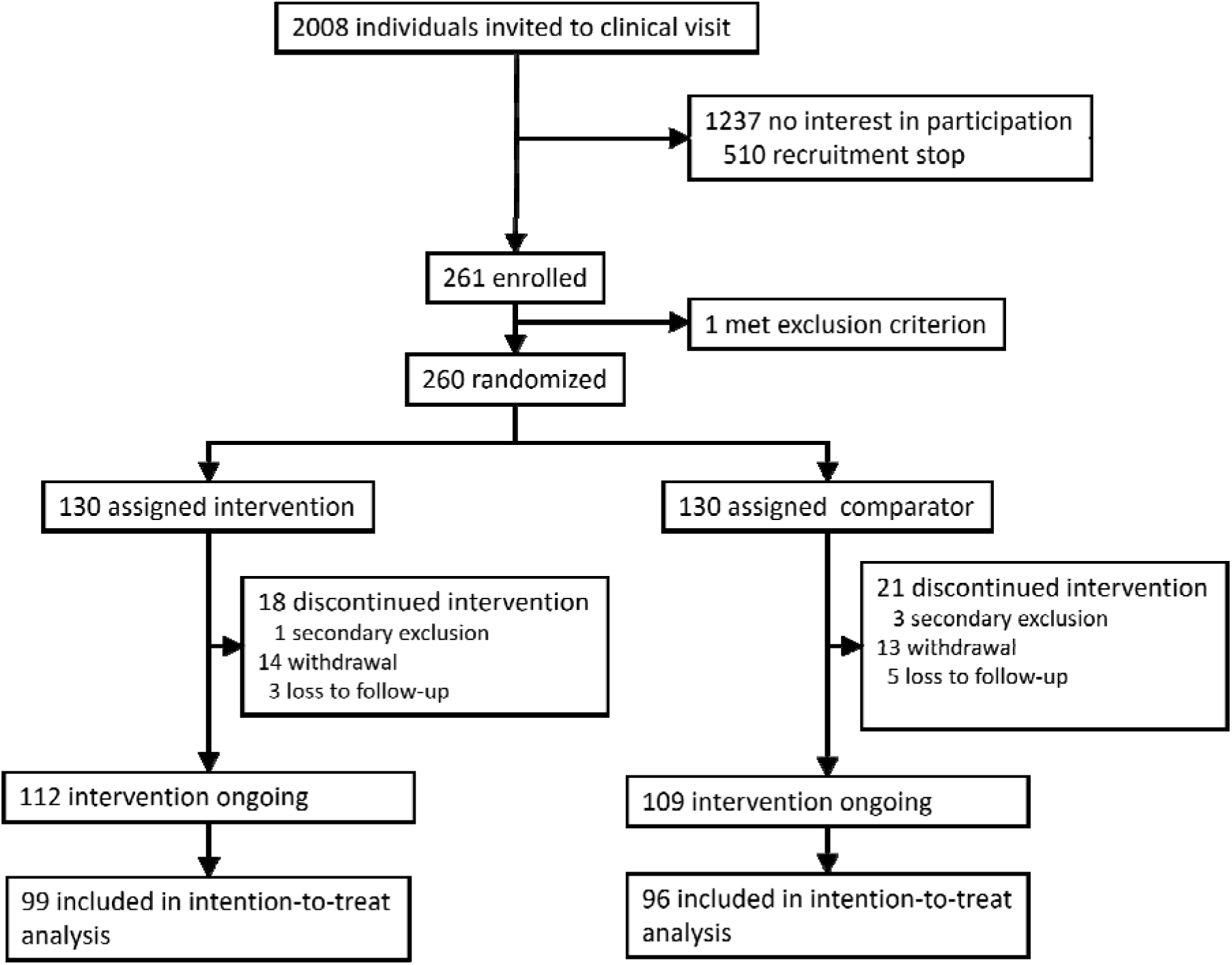
Participant flow.

**Table 1:** Distribution of demographics variables.

| Characteristics | Complete data | Intervention arm | Comparator arm |
| --- | --- | --- | --- |
| Female gender | 260 (100 %) | 60 (47 %) | 62 (47 %) |
| Completed weeks of pregnancy | 260 (100 %) | 39 (35-42) | 39 (34-42) |
| Birth weight in kg | 260 (100 %) | 3.4 (2.1-4.4) | 3.4 (1.8-4.5) |
| Cesarean section | 260 (100 %) | 50 (39 %) | 53 (40 %) |
| Maternal age at child's birth | 260 (100 %) | 34 (20-46) | 34.8 (21-47) |
| Maternal antibiotics during birth | 196 (75%) | 26 (27 %) | 27 (29 %) |
| Antibiotics after birth | 260 (100 %) | 6 (5 %) | 7 (5 %) |
| Breastfeeding duration in weeks | 194 (75%) | 34.4 (26-56) | 34.4 (23-52) |
| Secondhand Smoke | 243 (93%) | 14 (11 %) | 15 (12 %) |
| Presence of siblings | 260 (100 %) | 51 (40 %) | 59 (45 %) |
| Daycare attendance | 243 (93%) | 11 (9 %) | 12 (10 %) |
| Contact to animals in family | 243 (93%) | 46 (37 %) | 48 (40 %) |
| Parental asthma | 255 (98%) | 40 (32 %) | 42 (32 %) |
| Maternal asthma | 258 (99%) | 24 (18%) | 26 (20%) |
| Paternal asthma | 256 (98%) | 18 (15%) | 20 (15%) |
| Parental atopic eczema | 257 (99%) | 35 (28 %) | 38 (29 %) |
| Parental hay fever | 259 (100%) | 76 (59 %) | 78 (59 %) |
| Parental atopy | 250 (96%) | 89 (72 %) | 90 (71 %) |
| Parental milk allergy | 254 (98%) | 4 (3 %) | 3 (2 %) |
| Parents born abroad | 201 (77%) | 27 (26 %) | 36 (37 %) |
| Paternal school education $\geq 10$ years | 243 (93%) | 112 (91 %) | 116 (96 %) |
| Maternal school education $\geq 10$ years | 243 (93%) | 118 (96 %) | 119 (99 %) |
| Rural residential area | 243 (93%) | 27 (22 %) | 26 (21 %) |
| Cow's milk during pregnancy | 260 (100 %) | 121 (94 %) | 128 (97 %) |
| Cow's milk before intervention | 260 (100 %) | 64 (50 %) | 64 (49 %) |
| Infant formula before intervention | 260 (100 %) | 86 (67 %) | 86 (66 %) |
| Atopic eczema before intervention | 247 (95%) | 5 (4 %) | 6 (5 %) |

Complete data refers to absolute numbers. Frequencies and percentages per study arm are weighted and rounded to integers, which explains deviation from the original distribution of 130 individuals per study arm. Unweighted baseline data are published elsewhere.<sup>15</sup>

In the intervention group, 28 of 99 individuals (weighted: 38 of 130) were positive for the primary outcome atopic sensitization with any sIgE≥0.7 kU/l, and 24 of 96 in the comparator group (weighted: 33 of 130; RR= 1.13 [0.67 - 1.89]). While the intervention was unrelated to sIgE≥0.7 kU/l for any subsets of allergens (Supplemental Table 2), low-grade sensitization (sIgE≥0.35 kU/l) to milk allergens was more common in the intervention arm (RR=3.02 [1.27-7.19]), particularly for the allergens β-lactoglobulin and F2, which among others includes bovine serum albumin (BSA, Figure 2).

**Figure 2:**
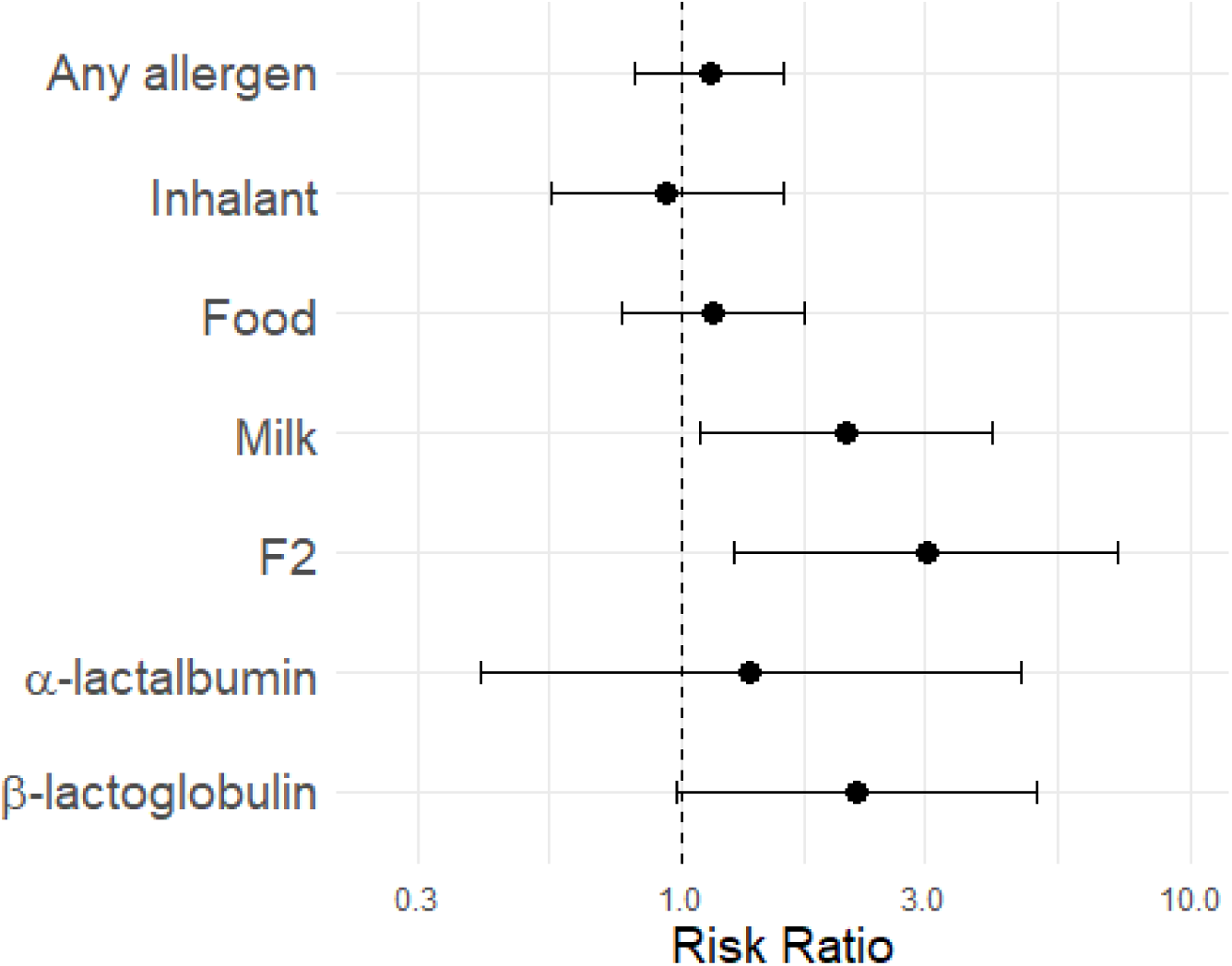
Low-grade atopic sensitization (sIgE ≥ 0.35 kU/l) after the intervention. Association of intervention and sensitization at CAP class 1 after completion of the intervention. Shown are relative risks (RR) with 95% confidence intervals. All analyses are weighted by inverse probability of study arm membership. F2 is a combination of α-lactalbumin, β-lactoglobulin, and bovine serum albumin (BSA).

For onset of asthma, 14 of the 112 individuals in the intervention group and 17 of the 109 individuals in the comparator group experienced an event (HR=0.55 [0.27−1.16], Figure 3A, Supplemental Table 3). This association was stronger and nominally significant in the 82 children with parental asthma (4 vs. 10 cases; post-hoc analysis: HR=0.30 [0.09−0.93], Figure 3B). It was not confounded by adherence to milk consumption as the latter was unrelated to parental asthma (Supplemental Figure 2). In a retrospective survey after completion of the intervention, 66 of 167 responding families incorrectly guessed the study arm they belonged to, thereby indicating maintained blinding. In these 66 participants, the point estimate remained inverse albeit with a larger confidence interval (HR=0.31 [0.06-1.63]).

**Figure 3:**
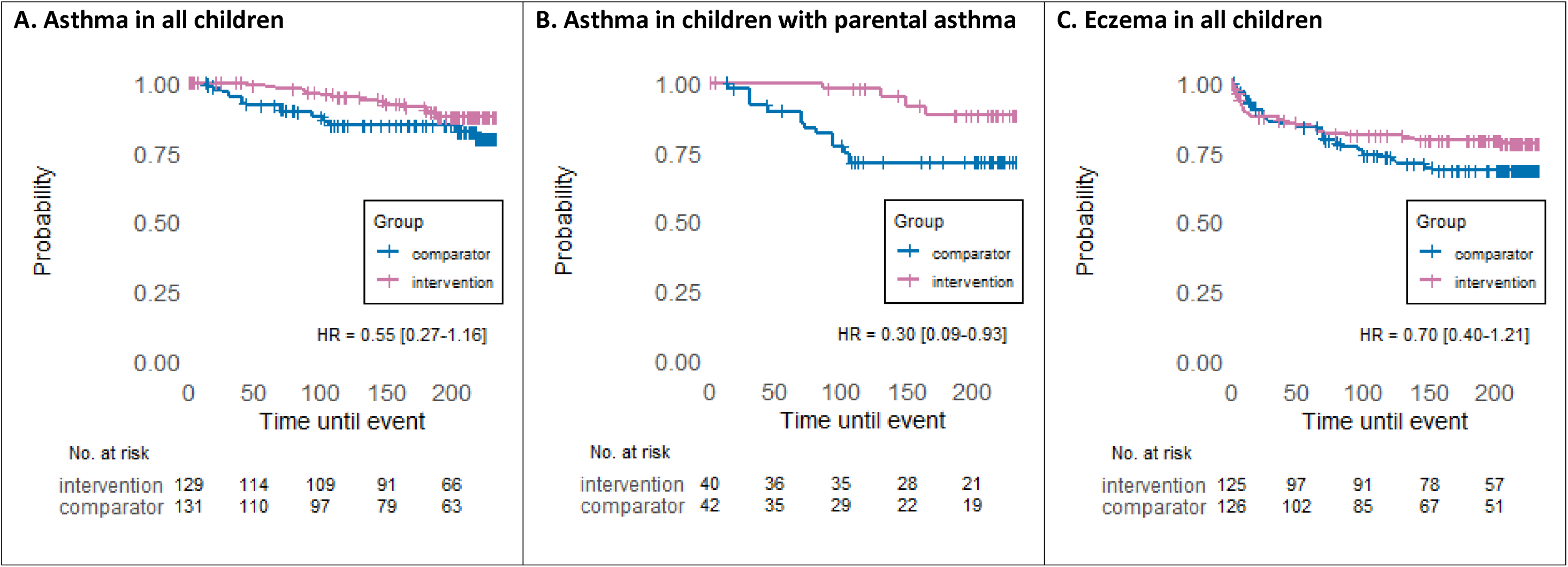
Time to onset of asthma and eczema stratified by study arm. All panels report weighted Kaplan-Meier estimates for time to onset of asthma (A,B) or eczema (C) stratified by study arm (intervention versus comparator group). Time is given in weeks from start of intervention. Hazard ratios (HR) are given with 95% confidence intervals and p-values derived from weighted survey Cox models. All analyses are weighted by inverse probability of study arm membership.

For onset of atopic eczema, 26 individuals of the intervention group and 33 individuals of the comparator group experienced an event (HR=0.70 [0.40−1.21], Figure 3C). After 65 weeks of age, events occurred less frequently in the intervention arm (post-hoc analysis: HR=0.22 [0.08−0.62], Figure 3C).

Low grade inflammation as determined by hs-CRP was not associated with the intervention (weighted geometric means ratio= 0.75 [0.35-1.60]).

The secondary endpoints related to respiratory tract infections were collected on a weekly basis and were rather uncommon during summer and winter holiday seasons and during the three intercurrent lockdown phases, which were followed by rebound effects, particularly in fall 2021 (Figure 4A). These rebounds affected the children differently, as illustrated by distinct trajectories of rhinitis in children born before and after the median birthdate (Supplemental Figure 3). Inverse associations of the intervention with rhinitis and cough were seen in the older half of the population (RR=0.82 [0.67-1.00] and RR=0.75 [0.60-0.94], respectively) but were lost in the younger half (Figure 4B/C, Supplemental Table 4).

**Figure 4:**
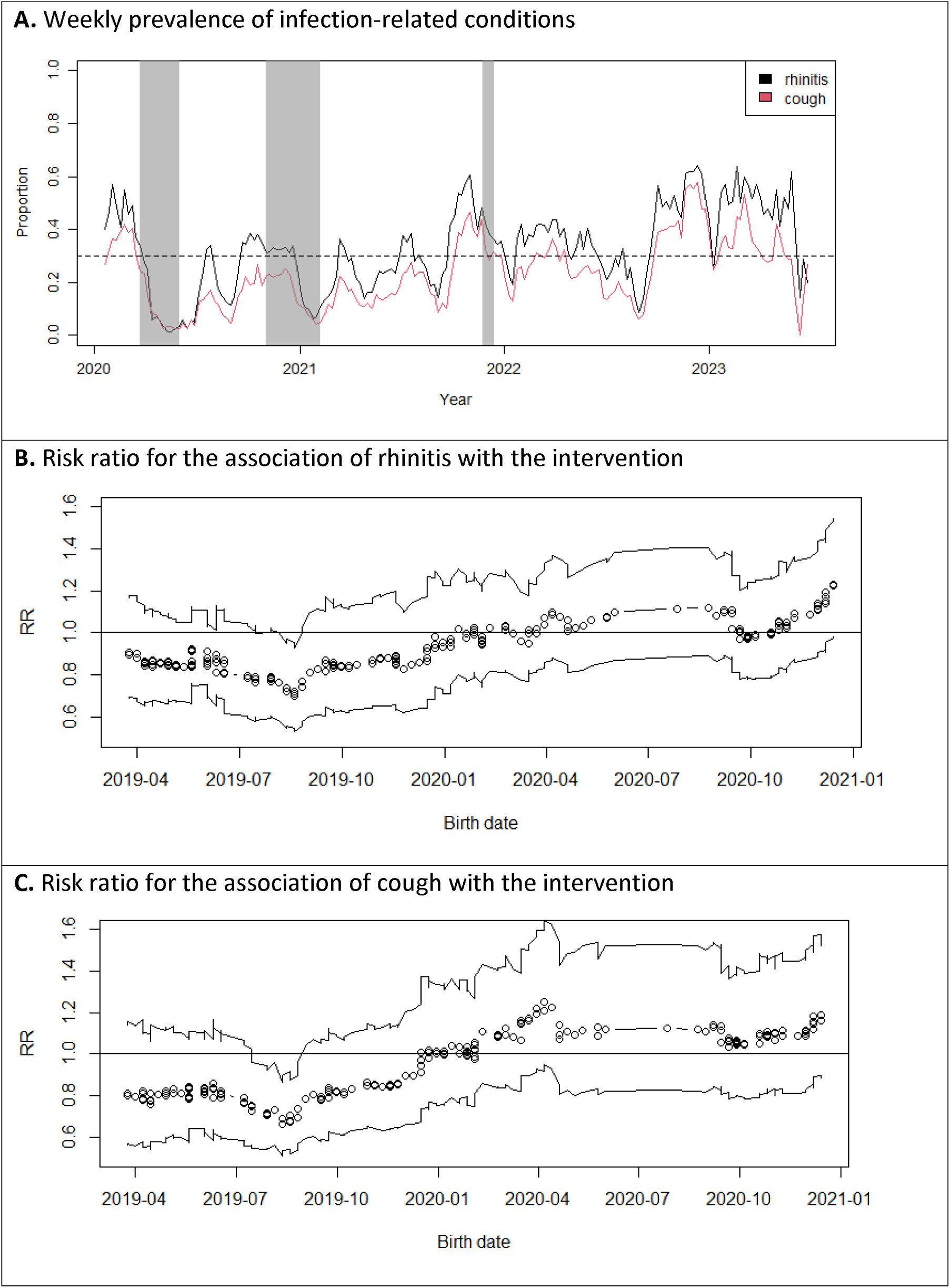
Respiratory tract infections over time. A. The shaded areas denote the lockdown periods in Bavaria, Germany. The horizontal line marks a weekly prevalence of 30%. B/C. Ninety weeks (42%) with a rhinitis prevalence below 30% are excluded.

As a biological marker of adherence to the protocol with respect to milk consumption, we assessed the fatty acid composition in erythrocyte membranes. The concentrations of the n-3 polyunsaturated fatty acids alpha-linolenic acid (ALA, C18:3-n3) and eicosapentaenoic acid (EPA, C20:5-n3) were higher in the intervention as compared to the comparator milk (Figure 5A). In erythrocyte membranes, EPA and docosapentaenoic acid (DPA, C22:5-n3) were higher in the intervention arm (Figure 5B).

**Figure 5:**
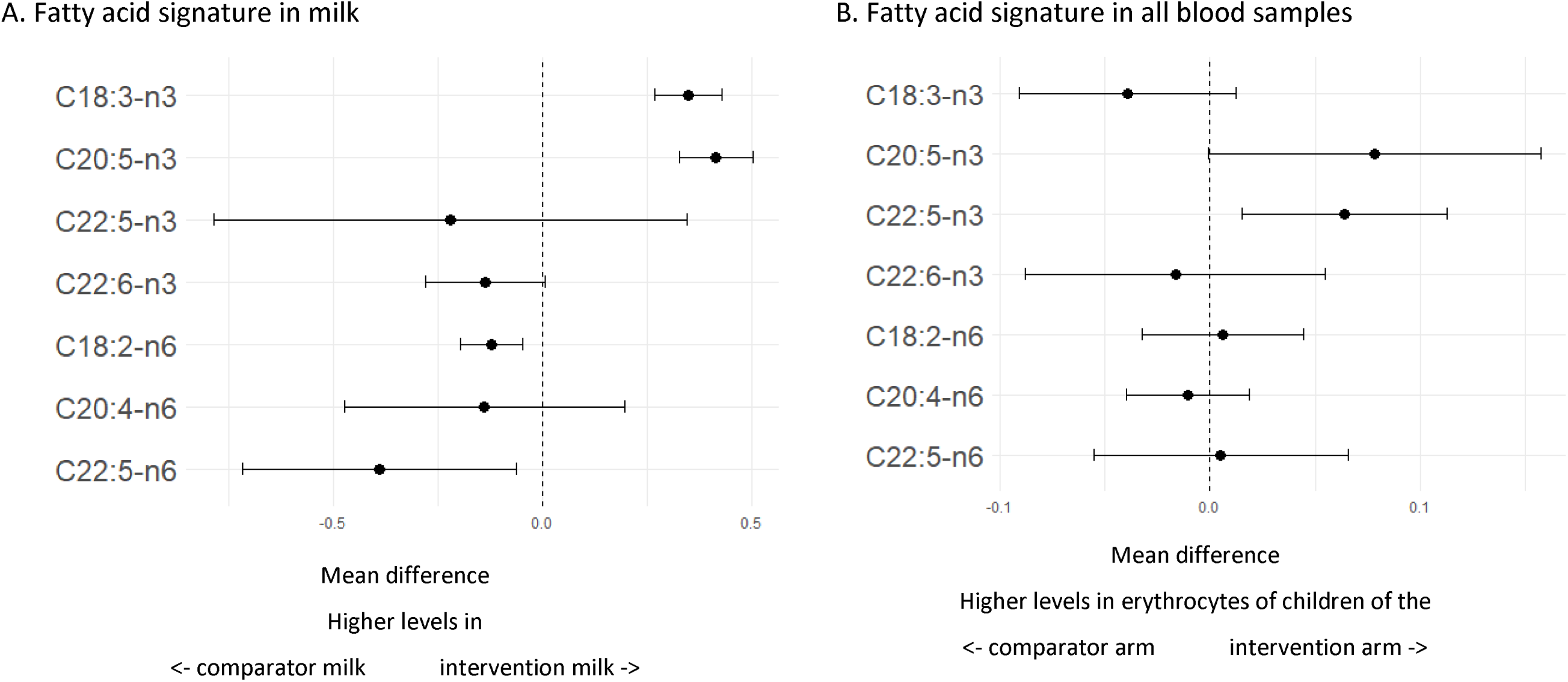
Fatty acid signatures in intervention milk and erythrocyte membranes. A. Fatty acid concentrations in relation to milk type (intervention versus comparator). Measurements in both milk types were performed in triplicates. B. Fatty acid concentrations in relation to study arm (intervention versus comparator) in all 198 children with measurements during the intervention. Both panels show beta estimates with 95% confidence intervals from unweighted (A) or weighted (B) linear regression

In children with parental asthma, the butyrate-producer maturation score was associated with the intervention and inversely with asthma (Figure 6).

**Figure 6:**
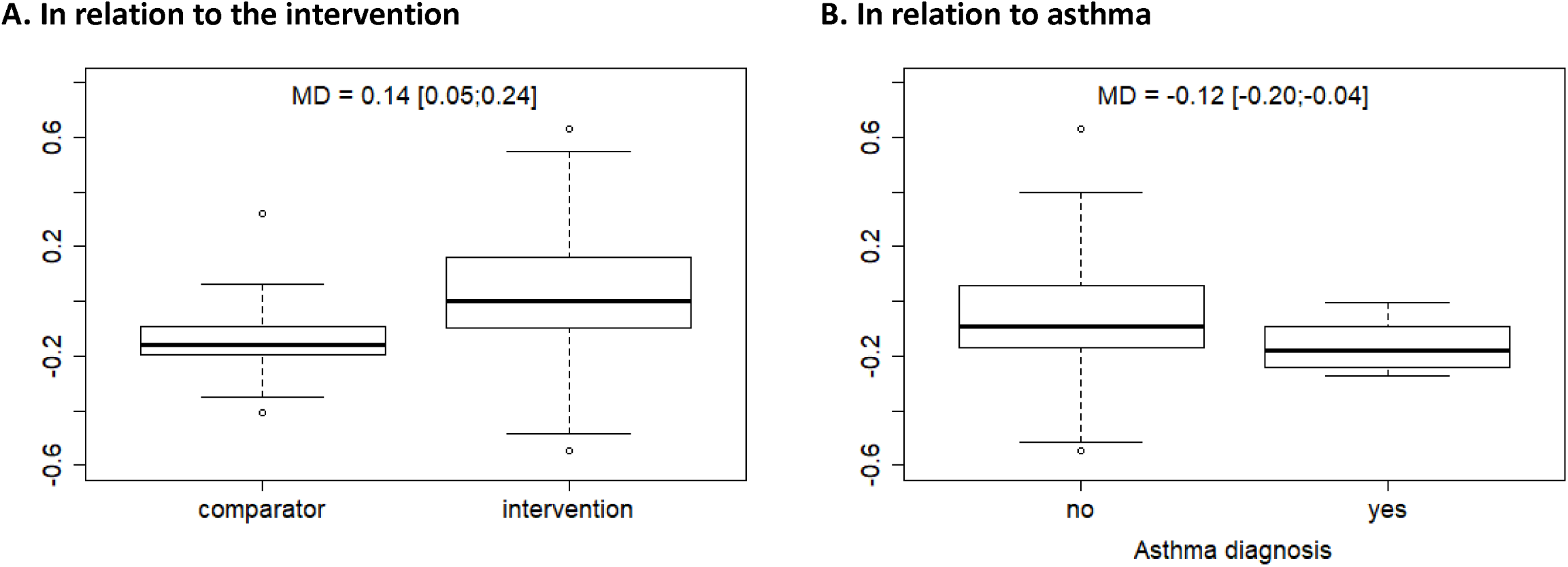
The butyrate-producer maturation score in children with parental asthma. The butyrate-producer maturation score was calculated in the PASTURE study according to Depner et al 2020^18^ and predicted in MARTHA samples. The butyrate-producer maturation score was calculated as the difference between predicted values of the butyrate-producer score at 12 months and at enrolment. A. Association with the intervention. B. Association with asthma. Restricted to children with parental asthma (n=68). All analyses are weighted by inverse probability of study arm membership. Mean differences (MD) are shown with 95% confidence intervals.

Adverse events and serious adverse events have been reported previously.^15^ Because of symptoms suggestive of milk allergy or intolerance, cow’s milk consumption was discontinued in 42 individuals, but symptoms did not resume after reintroduction of cow’s milk in all but one child. In the latter, milk allergy was excluded by a food challenge test under clinical conditions. Serious adverse events were reported for one child in the intervention and five children in the comparator arm and included failure to thrive (n=2), hospitalization for COVID19 (n=1), and unscheduled hospitalization over more than three days (n=3). All SAEs were unrelated to milk consumption as decided by the clinical investigators.

The difference in fat content of the intervention (4% fat) and comparator milk (1.5%) was unrelated to average body weight during and after the intervention (Supplemental Table 4).

## Discussion

The primary endpoint of the MARTHA study, atopic sensitization to any allergen, was unrelated to the intervention with minimally processed full-cream milk. However, the intervention showed a significant inverse association with the secondary outcome asthma in children with parental asthma. Likewise, the intervention showed an inverse association with respiratory tract infections represented by rhinitis and cough albeit only in the older half of the children. For atopic eczema, an inverse association was seen for a late-onset form.

Atopic sensitization as the primary endpoint was unrelated to the intervention, and a reasonable explanation is currently missing. However, the intervention was positively associated with low-grade sensitization to milk allergens, particularly β-lactoglobulin (BLG) and possibly BSA, albeit in the absence of clinical milk allergy.^15^ This seemingly paradoxical phenomenon was anticipated from earlier findings in the PASTURE study: Low-grade sensitization to cow’s milk was more common in farm children and inversely related to a family history of atopy, thereby suggesting a transient non-atopic, probably physiological IgE production without clinical relevance.^19,20^

Since BLG and BSA have previously been inversely associated with childhood asthma,^9^ induction of asymptomatic sIgE production by both allergens as shown here suggests that both whey proteins are functional and possibly involved in the effect of minimally-processed milk. While the comparator milk was treated by UHT, which inactivates most immunoactive whey proteins, the intervention milk underwent simple pasteurization thereby preserving whey proteins such as xanthine dehydrogenase, lactoferrin, serotransferrin, and polymeric immunoglobulin.^21^ Denaturation of β-lactoglobulin is reversible below 75°C,^22^ and minimal processing does not exceed this limit. Our intervention milk contained 30% BSA and 84% BLG in their native form compared to raw milk, while the comparator contained only 1% and 18% in native form, respectively (Supplemental Table 6).

Other whey proteins are more sensitive to heat treatment and are more extensively denatured in minimally-processed milk. However, heat inactivation seems to be inevitable as illustrated by recent outbreaks of milk-borne infections in California^23^ and issues of emerging antimicrobial resistance in raw milk farming.^24^

Besides intensity of heat treatment, the intervention milk differed from the comparator milk with respect to fat fraction and absence of homogenization.^15^ The latter disrupts fat globule membranes and renders triglycerides exposed to enzymatic alterations.^11^ Therefore, we hypothesized that the different milk types influenced the fatty acid composition of erythrocyte membranes, which can serve as longtime memory of fatty acid consumption. We found higher levels of the C18 and C20 n-3 polyunsaturated fatty acids in the intervention than in the comparator milk. The n-3 variants of these fatty acids are metabolized towards anti-inflammatory mediators, while the n-6 variants lead to pro-inflammatory mediators.^25^ The C18 and C20 fatty acids can be enzymatically elongated to C20 and C22, respectively, involving the fatty acid desaturase synthetases (FADS1/2), whose relevance in childhood asthma has repeatedly been discussed.^26^ The C20 and C22 n-3 fatty acids were detected at higher levels in erythrocyte membranes of children in the intervention group supporting adherence to milk consumption.

Until 60 weeks of age the proportion of individuals consuming the entire daily dose gradually increased from less than 20% to about 40% (Supplemental Figure 2). This protracted increase in adherence might reflect delayed weaning and may have resulted in an absent association with atopic eczema until age 65 weeks. Alternatively, the intervention may only affect a milder late-onset trajectory of eczema.^27,28^

The original primary aim of MARTHA was to demonstrate an effect of the intervention on asthma but was abandoned for the large sample size needed. We were surprised to find a significant inverse association with asthma in the 82 children with parental asthma. As the stratification by family history was only a proxy for the prespecified stratification for polymorphisms in the FADS1/2 genes, this finding must be interpreted cautiously.

Nevertheless, the restriction of inverse associations to children with parental asthma parallels findings from studies on atopic eczema, where the effect was similarly linked to a positive family history.^29,30^

Parental asthma may stand for an (epi-)genetic background with high penetrance.^31^ Polymorphisms or epigenetic modifications of genes encoding enzymes of the fatty acid metabolisms such as the above-mentioned FADS1/2 may be involved.^32–34^ A study on supplementation with fish oil during pregnancy showed a greater effect on subsequent asthma in the offspring of mothers with a FADS genotype associated with low EPA and DHA blood levels.^35^

In children with parental asthma, the butyrate-producer maturation score was associated separately with intervention and asthma status. This parallels findings from the PASTURE study, where a mature composition rich in butyrate-producing bacteria was associated with raw milk consumption and a lower risk of asthma.^18^ Butyrate is produced by fermenters in the colon and sensed by the peroxisome proliferator-activated receptor gamma (PPARγ) in colonocytes, which provide an anaerobic milieu in the gut, foster other beneficial fermenters, and prevent dysbiosis.^36^ In addition, butyrate inhibits inflammation through other mechanisms along the gut-lung axis.^36,37^ The association of the intervention with asthma emerged already before age 3 years, thereby suggesting a particular association with early, virus-triggered forms of asthma. This is also supported by the relatively high proportion of asthma cases (74%) defined by recurrent bronchitis. In addition, the consistent associations with infections of the upper and lower airways as illustrated by rhinitis and cough point towards an antiviral function of the intervention, at least in the older half of the children. Likewise, raw milk was inversely related to respiratory infections early in life in PASTURE.^12^

Although random variation cannot be excluded as an explanation of the abrupt loss of the inverse association of the intervention with rhinitis and cough in younger children, the effect modification by birthdate seems plausible given the more aggravated clinical presentations of viral infections particularly in young children in the wake of the first lockdown measures.^38–40^ The older half of the MARTHA participants might have already profited from the intervention in their first two years, i.e. before the intensive infection season in fall/winter 2021/2022 (Figure 4A).

A major strength of the MARTHA study is the randomized interventional design in a field largely dominated by observational evidence.^11^ The study is further strengthened by longitudinal follow-up, repeated symptom reporting on a weekly basis, biological exposure markers based on fatty-acid composition measured in erythrocyte membranes, and ancillary microbiome analyses providing mechanistic clues. The MARTHA trial was inspired by the PASTURE birth cohort, which enhances comparability and cross-validation of observational and interventional data.

The most obvious limitation of the MARTHA study is its continuation as a non-confirmatory trial, which disallows conclusive evidence. Nevertheless, the current results support the usefulness of a definitive trial. Since the association with asthma was concentrated on children with a positive family history, it may help finding a prevention strategy tailored to individuals at the highest risk of asthma.

Given the reduced sample size and exploratory nature of the analyses, adjustment for multiple testing was not applied. Accordingly, nominally significant associations should be interpreted as hypothesis-supporting rather than confirmatory.

While lung function testing cannot be performed in very young children, the outcome definition of parent-reported diagnosis of asthma or recurrent obstructive bronchitis is sensitive to accidental unblinding of parents, e.g. by recognizing non-homogenized milk by creaming. However, the inverse association of intervention and asthma was largely preserved in the unblinded population. A definitive trial will require a longer follow-up time as was possible in the MARTHA trial and should involve more objective criteria such as lung function measures. As all previous observational studies on the farm milk effect used the ISAAC definition of asthma, its application might be justifiable in this first interventional approach.

A minor limitation was the chance imbalance in measured baseline covariables following randomization, which was substantially mitigated by inverse probability of treatment weighting.

The absence of intervention-related adverse events was reassuring,^15^ although the sample was insufficient to establish uncommon harms. Minimally-processed, pasteurized milk is a microbiologically safer alternative to raw milk and may offer some potential for beneficial health effects as suggested by these preliminary data.

In conclusion, the prematurely terminated MARTHA trial did not demonstrate a reduction in the amended primary outcome of atopic sensitization. Findings for the composite asthma/recurrent-bronchitis outcome, eczema, respiratory symptoms and biological markers arose from multiple largely exploratory analyses and require confirmation in a prospective definitive trial.

## Other Information

### Registration

The Martha trial was registered on 2018-10-01 under the trial registration number DRKS00014781 (Universal Trial Number: U1111-1216-7580) with the German registry for clinical studies (DRKS, https://www.drks.de/search/de/trial/DRKS00014781/details).

### Protocol

The protocol can be obtained from the clinical registry.

### Role of the funding agency

The funders did not have a role in design, conduct, analysis, and reporting of the MARTHA trial.

### Data sharing

The R code of the original analysis can be found in the supplemental material. Upon application, the data can be analyzed on a protected platform (https://trace.ibe.med.uni-muenchen.de/). DNA sequences from fecal samples can be accessed under the accession number PRJEB127802 via https://www.ebi.ac.uk/ena/browser/home.

### Patient/public involvement

The study was mainly funded by a patient organization (Dutch Longfonds), and their patient advisory committee (PAC) helped with sharing patients’ perspectives and creating questionnaire items in easy language. Representatives of the MARTHA trial met with PAC members on a bimonthly basis.

## Data Availability

The data can be analyzed on the protected platform TRACE upon application.The R code of the original analysis can be found in the supplemental material.

https://trace.ibe.med.uni-muenchen.de/

https://www.ebi.ac.uk/ena/browser/home

## Acknowledgements

We thank the participating families for their willingness to contribute to this trial and their strong adherence to the protocol. We acknowledge the excellent work of our clinical team (Irmgard Häuser, Dr. Stefanie Hollunder, Dr. Sheena Sharma, and Sibylle Contento). We are grateful to Dr. von Obernitz, Dr. Edler von Koch, Prof. Dr. Anthuber, Prof. Dr. Kabesch, and Prof. Dr. Mahner and their teams for their support with approaching parents to be in their birth clinics. We thank the patient advisory committee of Dutch Longfonds and the Deutscher Asthma- und Allergie-Bund (DAAB) for providing their perspective on patients’ needs and help with phrasing questionnaires. We thank the members of the data safety and monitoring committee: Prof. Dr. Michael Perkin, Prof. Dr. Joachim Heinrich, and Dr. Bernhard Haller. Finally, we are greatly indebted to Prof. Ulrich Mansmann, Director Emeritus of the Institute for Medical Information Processing, Biometry, and Epidemiology of LMU Munich, for biostatistical advice with planning and analyzing this study.

## Online Supplement

### Methods

#### DNA extraction and sequencing

DNA was extracted from 500µl homogenized stool sample using the phenol-chloroform DNA extraction protocol described by Lueders et al.^1^ Amplicon sequencing of the V4 hypervariable region of the 16S rRNA gene was performed on a MiSeq Illumina instrument (MiSeq Reagent Kit v3 (600 Cycle); Illumina, San Diego, CA, USA) using the universal eubacterial primers 515FB and 806RB,^2, 3^ extended with sequencing adapters to match Illumina indexing primers. To monitor potential contamination during DNA extraction and amplification, extraction blanks and PCR no-template controls were included throughout the workflow. PCR amplification was carried out using NEBNext high fidelity polymerase (New England Biolabs, Ipswich, USA) in a total volume of 25 µl containing 5 ng template DNA, 12.5 µL polymerase mix, 5 pmol of each primer, and 0.3% bovine serum albumin. Cycling conditions consisted of an initial denaturation at 98°C for 5 min, followed by 25 cycles of 98°C for 10 s, 55°C for 30 s, and 72°C for 30 s, with a final extension at 72°C for 10 min. Amplicons were purified using MagSi NGSprep Plus beads (Steinbrenner, Wiesenbach, Germany) and quantified via PicoGreen assay. Subsequently, indexing PCR was performed using the Nextera XT Index Kit v2 (Illumina, Inc. San Diego, CA, US) in a total volume of 25 µl (10 ng DNA template, 12.5 µl NEBNext high fidelity polymerase, 2.5 µl of each indexing primer) and the following PCR conditions: 30s at 98°C; 8 cycles of 10 s at 98°C, 30s at 55°C, 30s at 72°C; 5min 72°C. Indexed amplicons were purified using MagSi NGSprep Plus beads, qualified and quantified via a Fragment Analyzer™ instrument (Advanced Analytical Technologies, Inc., Ankeny, USA) and pooled at equimolar concentrations to 4nM prior to sequencing.

#### Bioinformatic processing

QIIME2 (v 2021.8) was used for the 16S amplicon analysis.^4^ Raw FASTQ files were generated as 300-bp paired-end reads and imported into QIIME 2 using the input format “PairedEndSequencesWithQuality”. Cutadapt was used to trim off the V4 primer.^5^ Paired-end 16S rRNA gene amplicon sequences were denoised, quality filtered, merged, and screened for chimeras using the DADA2 plugin in QIIME 2 (qiime dada2 denoise-paired).^6^ Forward and reverse reads were truncated based on quality profiles. Reads were additionally filtered using a quality score truncation threshold of Q2 (--p-trunc-q 2) and maximum expected error thresholds of 2 for forward reads and 4 for reverse reads. Error rates were learned from 1,000,000 reads (--p-n-reads-learn 1000000). The resulting amplicon sequence variants (ASVs) were inferred after denoising, paired-end read merging, and chimera removal. Taxonomic classification of amplicon sequence variants (ASVs) was performed in QIIME 2 using the feature-classifier plugin. A naïve Bayes classifier was trained on the SILVA v138 99% reference database after extracting the region corresponding to the amplified 16S rRNA gene fragment using the primer pair (forward: GTGYCAGCMGCCGCGGTAA; reverse: GGACTACNVGGGTWTCTAAT), which was used for amplification.^7^ Representative ASV sequences were then classified using the classify-sklearn method. The resulting taxonomic assignments were exported for downstream analyses. The decontam R package was used to identify and remove contaminants using the prevalence method.^8^ Eukaryota and Archea were excluded to only focus on Bacteria. Cyanobacteria, Chloroplasts and Mitochondria were also excluded a priori. Samples with < 1,000 reads were removed from the analysis.

#### Statistical analysis

For the butyrate-producer maturation score we reran the random forest analysis using the R package *ranger* for the butyrate score at 12 months in PASTURE, as described in Depner et al 2020.^9^ Thereby, we restricted the list of genera to those also detectable in MARTHA, i.e., *Anaerostipes, Blautia, Romboutsia, Faecalibacterium, Bifidobacterium, Intestinibacter, Roseburia, Turicibacter, Erysipelatoclostridium, Dorea, Lachnospiraceae* NK4A136 group*, Tyzzerella, Fusicatenibacter, Terrisporobacter, Lactobacillus, Lachnoclostridium, Sellimonas, Enterococcus, Akkermansia, Subdoligranulum, Clostridium* sensu stricto 1*, Agathobacter, Lactococcus, Actinomyces, Collinsella, Eggerthella, Streptococcus, Parabacteroides, Veillonella, Haemophilus, Bacteroides, Dialister, Alistipes, Prevotella, Holdemanella*. The prediction was dominated by *Blautia* (6.56), *Anaerostipes* (6.27), *Bifidobacterium* (5.79), *Faecalibacterium* (3.78), and *Romboutsia* (3.18), which showed variable importance factors (in brackets) greater than 3, while the other genera ranged between 0.01 and 1.04. Then the random forest prediction was applied to the MARTHA microbiome data at enrolment (on average at 9 months) and at 12 months of age. We calculated the butyrate-producer maturation score as the difference between the two time points, which yielded a normally distributed variable ranging from −0.54 to 0.70 with a mean of −0.02. In children with a family of asthma, we compared the butyrate-producer maturation score between the intervention and the comparator groups and between asthma cases and non-cases by a linear regression and derived the mean difference as the beta estimate.

## Supplemental Figures

**Supplemental Figure 1:**
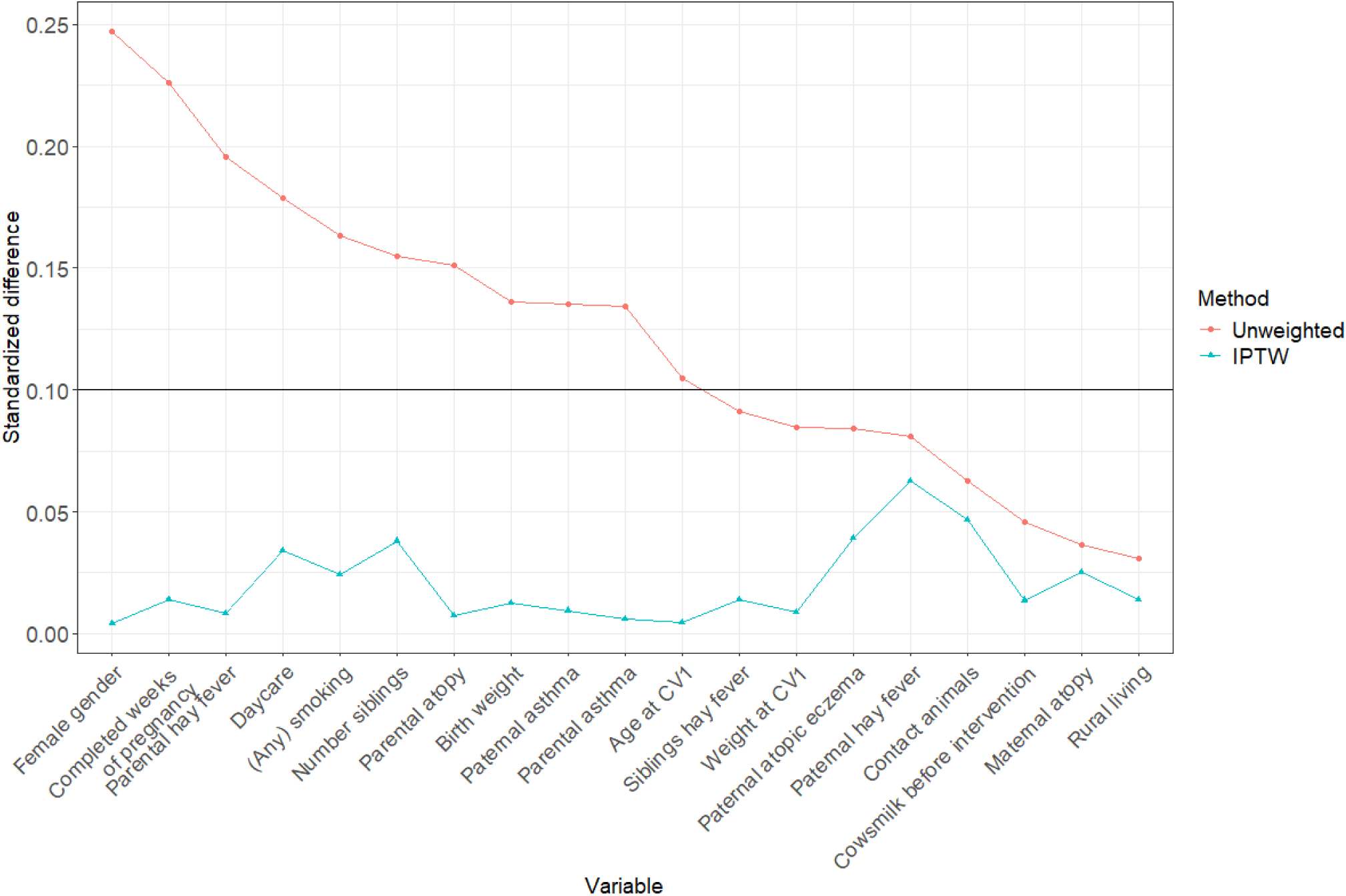
Inverse probability of treatment weighting. Standardized differences for all variables used for calculating the inverse probability of treatment weighting (IPTW) by a propensity score analysis for treatment allocation. The standardized differences using IPTW were all below 0.1 and thus considered acceptable, ultimately demonstrating equal distribution of variables when applying IPTW.

**Supplemental Figure 2:**
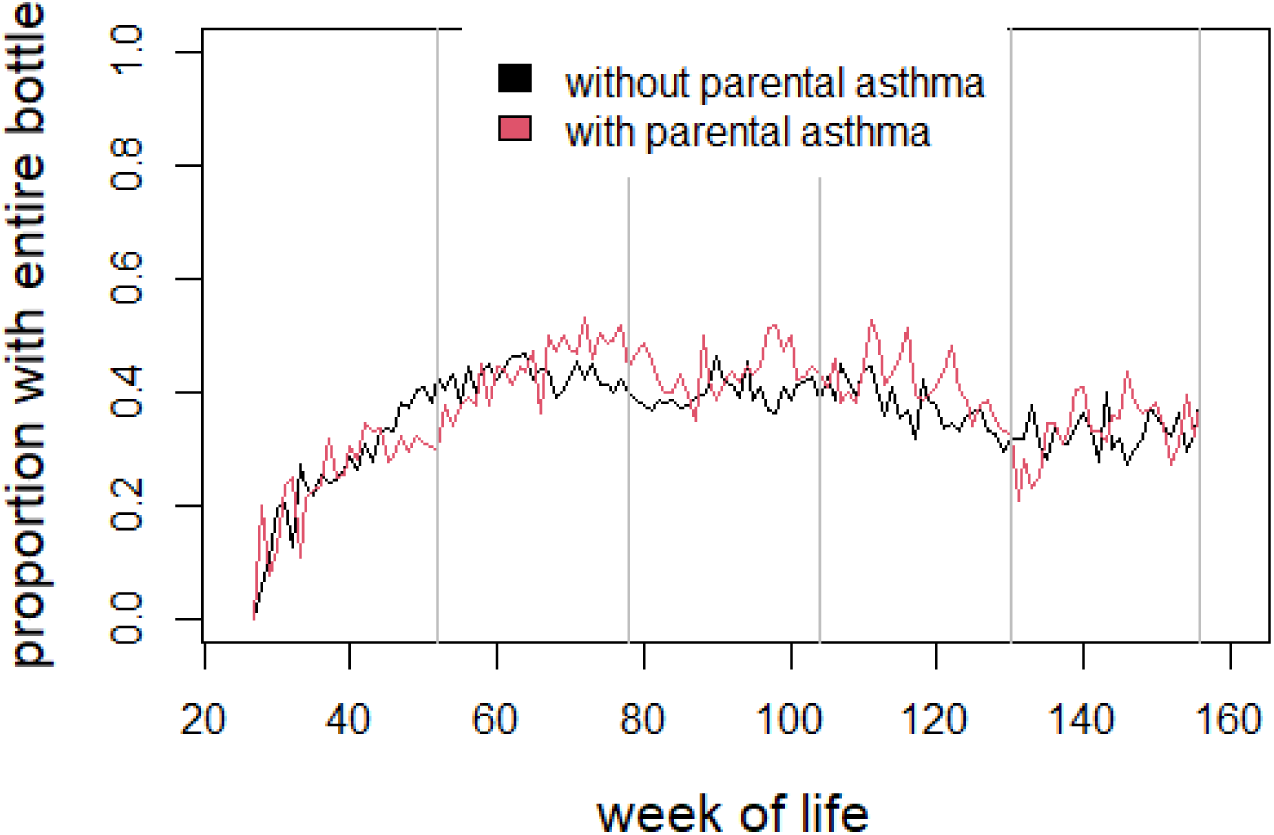
Adherence to milk consumption over age stratified by parental history of asthma. The proportion of children who drank the entire bottle on at least four days in the respective week is plotted against age in weeks. The analysis is weighted by inverse probability of study arm membership.

**Supplemental Figure 3:**
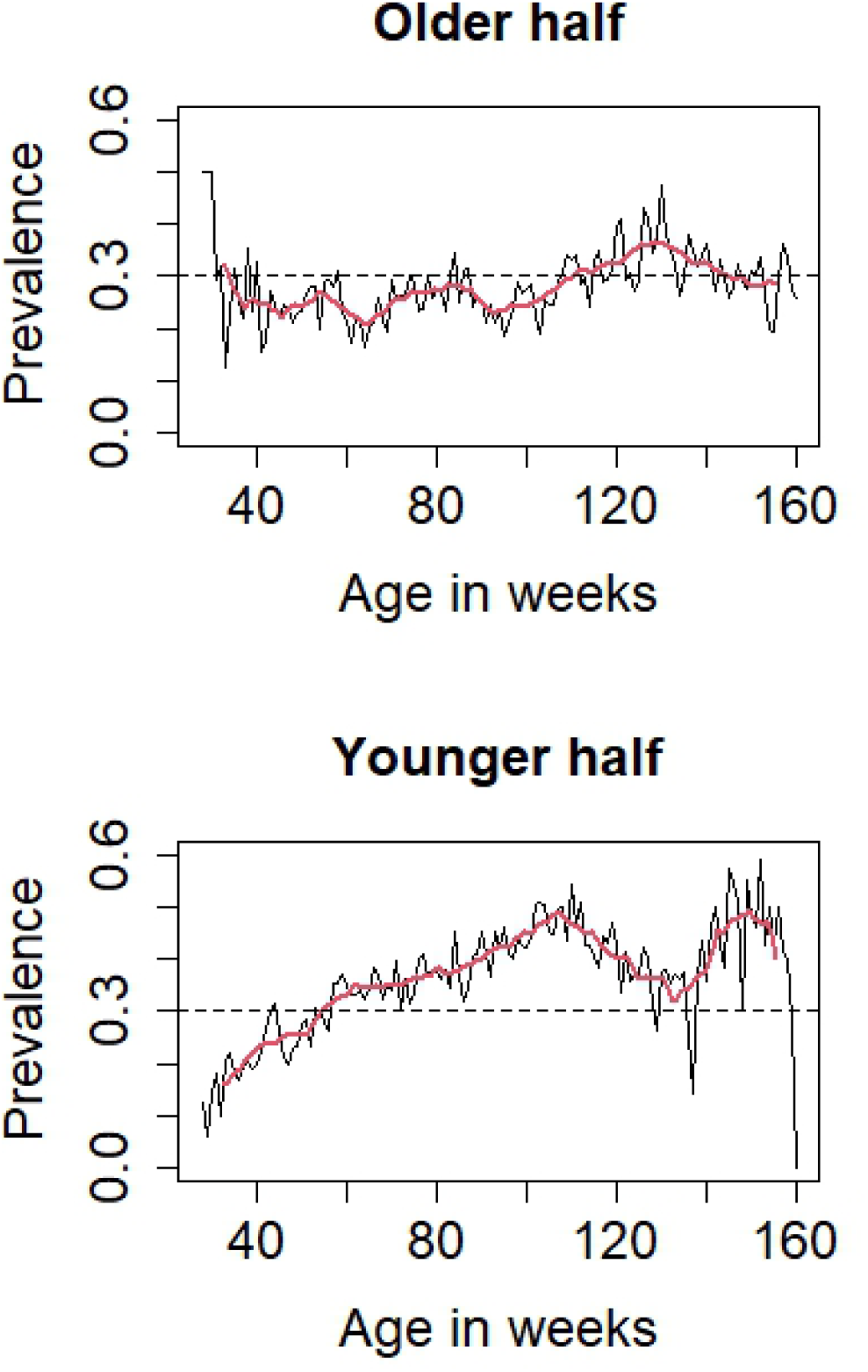
Associations of the intervention and rhinitis over age in relation to median of birth date. The black line shows the prevalence rhinitis per life week. The red line is a smoothed variant of the black line averageing over 10 weeks. The dashed horizontal line denotes a rhinitis prevalence of 30% per week. The median of birth dates was 2019-12-16.

## Supplemental Tables

**Supplemental Table 1:** Variables related to the intervention.

| Characteristics | Complete data | Intervention arm | Comparator arm |
| --- | --- | --- | --- |
| Age at clinical visit 1 in weeks | 260 (100 %) | 32.5 (22.6-60.6) | 33.4 (22.4-60.6) |
| Age at start of intervention in weeks | 257 (99%) | 35 (26-64) | 34.3 (26-61) |
| Age at end of intervention in weeks | 257 (99%) | 152 (32-156) | 148.8 (36-156) |
| Duration of intervention in weeks | 257 (99%) | 110 (1-130) | 107.1(2-130) |
| Entire serving on at least 365 days | 260 (100 %) | 36 (28 %) | 41 (31 %) |
| Additional milk consumption ≥25 weeks | 260 (100 %) | 67 (52 %) | 61 (46 %) |
| Weeks with additional milk consumption | 260 (100%) | 27.9 (9-35) | 26 (10-33) |

The analyses were performed using inverse probability of treatment weighting, and weighted estimates for frequencies were rounded to integer values.

**Supplemental Table 2:**
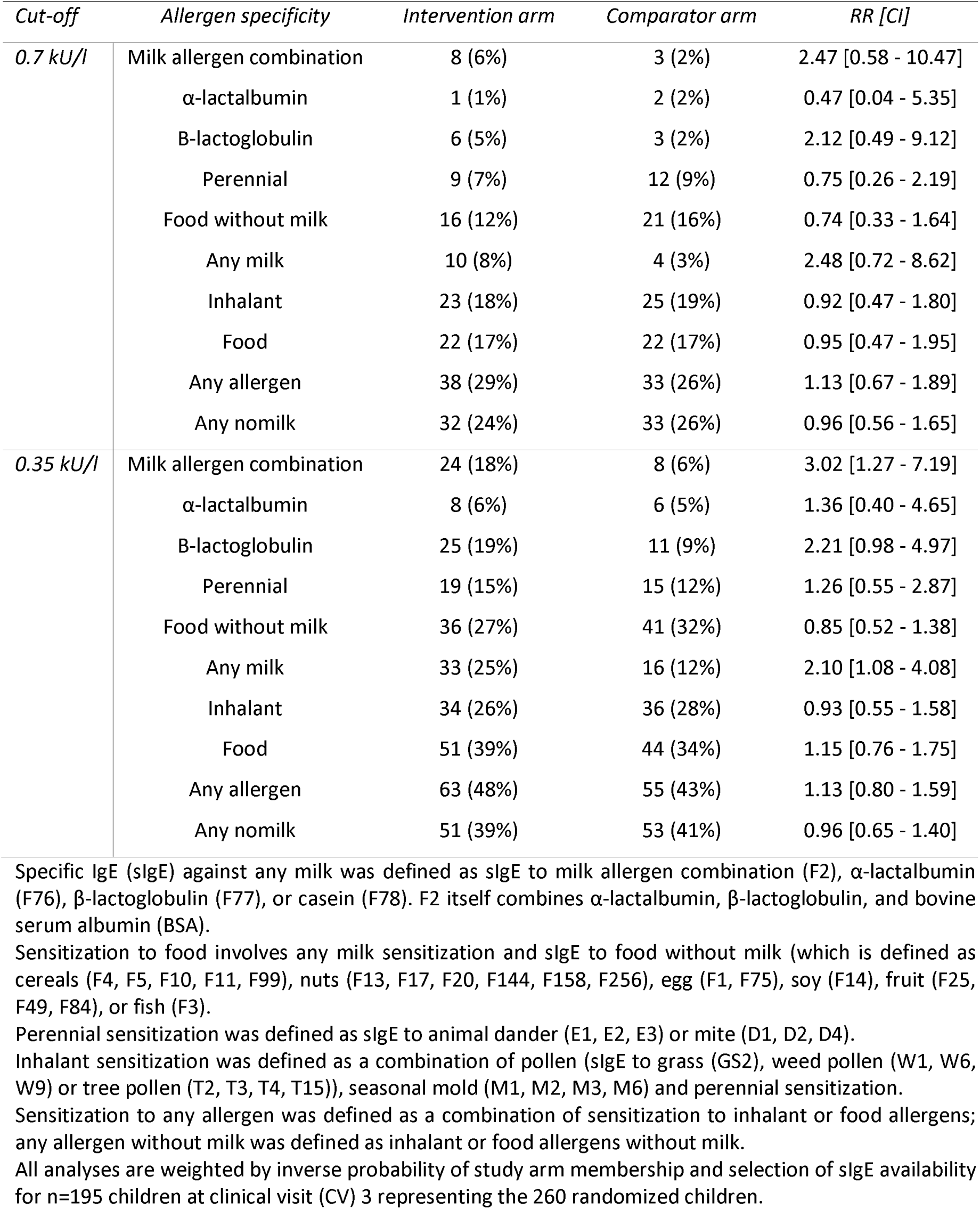
Prevalence of specific IgE by study arm at 3 years.

**Supplemental Table 3:** Description of asthma cases.

| Type of event | Study arm | Sex | Parental asthma | Number of cases |
| --- | --- | --- | --- | --- |
| Asthma diagnosis | Comparator | Male | No | 1 |
| Asthma diagnosis | Comparator | Male | Yes | 2 |
| Asthma diagnosis | Comparator | Female | No | 0 |
| Asthma diagnosis | Comparator | Female | Yes | 2 |
| Asthma diagnosis | Intervention | Male | No | 2 |
| Asthma diagnosis | Intervention | Male | Yes | 0 |
| Asthma diagnosis | Intervention | Female | No | 1 |
| Asthma diagnosis | Intervention | Female | Yes | 0 |
| Bronchitis twice | Comparator | Male | No | 3 |
| Bronchitis twice | Comparator | Male | Yes | 3 |
| Bronchitis twice | Comparator | Female | No | 3 |
| Bronchitis twice | Comparator | Female | Yes | 3 |
| Bronchitis twice | Intervention | Male | No | 3 |
| Bronchitis twice | Intervention | Male | Yes | 3 |
| Bronchitis twice | Intervention | Female | No | 4 |
| Bronchitis twice | Intervention | Female | Yes | 1 |

All 31 asthma cases are listed without any weighting.

**Supplemental Table 4:** Modification of the association of intervention and respiratory infections by a cohort effect.

| Outcome | Subset | Determinant | Risk ratio |
| --- | --- | --- | --- |
| **Rhinitis** | All | Intervention | 0.82 [0.67-0.997] |
|  |  | Younger than median | 0.99 [0.83-1.19] |
|  |  | Interaction term | 1.37 [1.06-1.76] |
|  | Younger than median | Intervention | 1.12 [0.95-1.31] |
|  | Older than median | Intervention | 0.82 [0.67-0.998] |
| **Cough** | All | Intervention | 0.75 [0.60-0.94] |
|  |  | Younger than median | 0.89 [0.72-1.09] |
|  |  | Interaction term | 1.54 [1.15-2.06] |
|  | Younger than median | Intervention | 1.15 [0.95-1.40] |
|  | Older than median | Intervention | 0.75 [0.60-0.94] |

Weighted risk ratios are given with 95%-confidence intervals. Only weeks with a rhinitis prevalence of at least 30% were included because of the overall low prevalence of respiratory infections during lockdown and holiday periods. The interaction term is a multiplicative term based on the two dichotomous variables “intervention” and “younger than median”. The median of birth dates was 2019-12-16.

**Supplemental Table 5:** Variables related to weight and height development.

| Characteristics | Complete data | Intervention arm | Comparator arm |
| --- | --- | --- | --- |
| Weight percentiles at CV1 | 260 (100%) | 54.2 (26.66) | 54.1 (28.50) |
| Weight percentiles at CV2 | 215 (83%) | 56.6 (28.89) | 58.4 (28.51) |
| Weight percentiles at CV3 | 193 (74%) | 52.0 (28.71) | 56.2 (27.06) |
| Height percentiles at CV1 | 260 (100%) | 42.8 (27.65) | 41.9 (30.09) |
| Height percentiles at CV2 | 216 (83%) | 45.9 (30.87) | 45.1 (30.40) |
| Height percentiles at CV3 | 193 (74%) | 51.6 (31.41) | 52.8 (29.17) |

For percentiles mean and standard deviation are given. The analyses were performed using inverse probability of treatment weighting. CV = clinical visit.

**Supplemental Table 6:** Protein contents in the intervention and comparator milks in relation to raw milk.

| **Protein** | **Intervention milk**<br>**compared to**<br>**raw milk (%)** | **Comparator milk**<br>**compared to**<br>**raw milk (%)** | **Comparator**<br>**compared to**<br>**intervention milk (%)** |
| --- | --- | --- | --- |
| Bovine Serum albumin (BSA) | 30 | 1 | 4 |
| Lactoferrin | 19 | 1 | 5 |
| Ig-like domain-containing protein | 99 | 7 | 7 |
| Polymeric immunoglobulin receptor | 11 | 1 | 7 |
| Beta-lactoglobulin | 84 | 18 | 22 |
| Osteopontin | 16 | 5 | 30 |
| Alpha-lactalbumin | 59 | 23 | 39 |

The intervention and comparator milks (single measurements) are compared to the mean of raw milk measurements from three different farms. In the left two columns, the protein concentration in raw milk is set to 100%. In the rightmost column, the protein concentration in the intervention milk is set to 100%.

