## Supplementary material for "Minimally processed cow’s milk and early childhood asthma-related outcomes: exploratory findings from the early terminated MARTHA randomized trial": R-Code

```
# MARTHA trial
# Functions used to create svy tables
# 2022-01-04 meweber
# 2023-05-23 meweber: updated to provide group as an input to function

svychisqloop  <- function(vars ,design){svychisq(as.formula(paste0( "~" , vars)), design)}

# Descriptive table stratified for two groups with p-values - with complete cases
ta2.svy       <- function(xvar, yvar, dset, design) {
  y           <- dset[, yvar]
  cc      <- complete.cases(dset[, c(xvar, yvar)])
  n_cc    <- sum(cc)
  pct_cc  <- 100 * n_cc / nrow(dset)
  n           <- paste(yvar,": ",levels(y),sep="")

  fl          <- as.formula(paste0("~", yvar, "+", xvar))
  X           <- svytable(fl, design)
  if(dim(X)[1] == 1) {
    if(rownames(X) == "0") X <- rbind(X, c(0, 0)) else X <- rbind(c(0, 0), X)
    rownames(X)  <- c("0", "1")}
  fm          <- paste0(yvar, "+", xvar)

  R           <- matrix("" ,nrow=max(2,dim(X)[1]), ncol=9)
  R[1,8]     <- n_cc
  R[1,9]     <- paste(round(pct_cc), "%")
  R[,1]       <- n
  R[,2]       <- round(X[,"1"])
  R[,3]       <- paste(round(100*X[,"1"]/sum(X[,"1"])),"%")
  R[,4]       <- paste(round(100*X[,"1"]/rowSums(X)),"%")
  R[,5]       <- round(X[,"0"])
  R[,6]       <- paste(round(100*X[,"0"]/sum(X[,"0"])),"%")
  R[,7]       <- paste(round(100*X[,"0"]/rowSums(X)),"%")
  colnames(R) <- c("Variable", "Test.milk_n","Test.milk_pct","Test.milk_rowpct",
                   "Control.milk_n","Control.milk_pct","Control.milk_rowpct",
                   "Complete_n","Complete_pct")
  return(data.frame(R,stringsAsFactors = FALSE))
}

TA.svy        <- function(group, var.list, dset, design) {
  if (length(table(dset[,group]))==2) {
  TA        <- NULL
    for (var in var.list) TA <- rbind(TA,ta2.svy(xvar = group, yvar = var, dset, design))
    return(TA)
  }
}

# with complete cases
num.svy       <- function(var, xvar, dset, name, design, digits=1){
  cc          <- complete.cases(dset[, c(var, xvar)])
  n_cc        <- sum(cc)
  pct_cc      <- round(100 * n_cc / nrow(dset), 1)
  cc_out      <- paste0(n_cc, " (", pct_cc, "%)")

  g1          <- round(summary(dset[, var][dset[, xvar]==1]),digits=digits)
  g0          <- round(summary(dset[, var][dset[, xvar]==0]),digits=digits)

  var1        <- as.formula(paste0("~",var))

  gs1         <- oldsvyquantile(var1, design = subset(design, eval(parse(text =
paste0("dset$", xvar))) == "1"), quantile = c(0.25, 0.5, 0.75), na.rm = TRUE)
  gs1         <- round(gs1[,2], digits = digits)

  gs0         <- oldsvyquantile(var1, design = subset(design, eval(parse(text =
paste0("dset$", xvar))) == "0"), quantile = c(0.25, 0.5, 0.75), na.rm = TRUE)
  gs0         <- round(gs0[,2], digits = digits)

  g1          <- ifelse(var %in% quartile, paste0(gs1," (",round(g1["1st Qu."]),"-
",round(g1["3rd Qu."]),")"), paste0(gs1," (",g1["Min."],"-",g1["Max."],")"))
```

```
  g0          <- ifelse(var %in% quartile, paste0(gs0," (",round(g0["1st Qu."]),"-
",round(g0["3rd Qu."]),")"), paste0(gs0," (",g0["Min."],"-",g0["Max."],")"))
  return(c(name, cc_out, g1, g0))
}

tab.num.svy   <- function(x, group, dset, name, design) {
  num.svy(var =  x, xvar = group, dset, name = name$N[name$V == x], design = design, digits =
1)
}

# for percentiles (giving mean and standard deviation) - with complete cases
num.svy.mean       <- function(var, xvar, dset, name, design, digits=1){
  cc          <- complete.cases(dset[, c(var, xvar)])
  n_cc        <- sum(cc)
  pct_cc      <- round(100 * n_cc / nrow(dset), 1)
  cc_out      <- paste0(n_cc, " (", pct_cc, "%)")

  g1          <- round(summary(dset[, var][dset[, xvar]==1]),digits=digits)
  g0          <- round(summary(dset[, var][dset[, xvar]==0]),digits=digits)

  var1        <- as.formula(paste0("~",var))

  gs1         <- svymean(var1, design = subset(design, eval(parse(text =  paste0("dset$",
xvar))) == "1"), na.rm = TRUE)
  gs1         <- round(gs1[[1]], digits = digits)

  gs0         <- svymean(var1, design = subset(design, eval(parse(text =  paste0("dset$",
xvar))) == "0"), na.rm = TRUE)
  gs0         <- round(gs0[[1]], digits = digits)

  g1          <- ifelse(var %in% quartile.mean, paste0(gs1," (",round(g1["1st Qu."]),"-
",round(g1["3rd Qu."]),")"), paste0(gs1," (", round(sd(dset[, var][dset[, xvar]==1], na.rm =
TRUE), 2),")"))
  g0          <- ifelse(var %in% quartile.mean, paste0(gs0," (",round(g0["1st Qu."]),"-
",round(g0["3rd Qu."]),")"), paste0(gs0," (", round(sd(dset[, var][dset[, xvar]==0], na.rm =
TRUE), 2),")"))
  return(c(name, cc_out, g1, g0))
}

tab.num.svy.mean   <- function(x, group, dset, name, design) {
  num.svy.mean(var =  x, xvar = group, dset, name = name$N[name$V == x], design = design,
digits = 1)
}

### eof
```

```
# MARTHA trial
# numbers for consort flow
# 2026 meweber

load(file.path(fp, "paper_outcomes_analysis_data.rdata"))
rm(per_milk)
rm(PM)

table(MC$verum)

consort           <- data.frame(intervention = c("130"), comparator = c("130"))

table(MC$type_termination, MC$verum, exclude = NULL)

consort           <- rbind(consort, c("18", "21"))
consort           <- rbind(consort, c("1", "3"))
consort           <- rbind(consort, c("14", "13"))
consort           <- rbind(consort, c("3", "5"))
consort           <- rbind(consort, c("112", "109"))

table(MC$in_IgE, MC$verum, exclude = NULL)

consort           <- rbind(consort, c("99", "96"))

rownames(consort) <- c("arm", "discontinued intervention", "secondary exclusion", "withdrawal",
"loss to follow-up", "intervention ongoing", "included in intention-to-treat analysis")

write.csv2(consort, file = file.path(op, paste("consort_flow.csv")))

rm(list = ls())

#### eof
```

```
# MARTHA trial
# Analysis IgE CV3 class 1
# 2024-12-04 meweber

library(survey)
library(ggplot2)
library(dplyr)
library(scales)

# load data set
load(file.path(fp, "paper_outcomes_analysis_data.rdata"))

rm(per_milk)
rm(PM)

MI                 <- subset(MC, !is.na(MC$ipw.IgE_sub))

# define survey design
ate                    <- svydesign(ids = ~ LID_trans, weights = ~ ipw.IgE_sub, dat = MI)

IgE.list.1             <- names(MI)[grep("cl1", names(MI))]

result_CV3_class_1 <- data.frame(NULL)
for(A in IgE.list.1) {
  fo                  <- as.formula(paste(A, "~ as.numeric(verum)"))
  model               <- svyglm(fo, design = ate, family = quasipoisson(link = "log"))
  RR                  <- round(exp(coef(model)[2]), 2)
  CI                  <- exp(confint(model)[2,])
  result_CV3_class_1 <- rbind(result_CV3_class_1, data.frame(A, summary(model)$coef[2,
c("Estimate")], summary(model)$coef[2, c("Pr(>|t|)")], RR, CI[1], CI[2], paste0(RR, " ", "[",
round(CI[1], 3), " - ", round(CI[2], 3), "]", ",", " ",
format(round(as.numeric(summary(model)$coef[2, "Pr(>|t|)"]), 3), scientific = FALSE))))
}

names(result_CV3_class_1)          <- c("Outcome", "Estimate", "p", "RR", "lowerCI", "upperCI",
"OR_CI_p")

# subset for allergens of interest
result_CV3_class_1_s <- subset(result_CV3_class_1, result_CV3_class_1$Outcome %in%
c("any_allergen_cl1", "food_cl1", "any_milk_cl1", "milk_cl1", "inhalant_cl1",
"a_lactalbumin_cl1", "b_lactoglobulin_cl1"))

## sort
result_CV3_class_1_s   <- result_CV3_class_1_s %>%
  mutate(Outcome = factor(Outcome, levels = c("any_allergen_cl1", "inhalant_cl1", "food_cl1",
"any_milk_cl1", "milk_cl1", "a_lactalbumin_cl1", "b_lactoglobulin_cl1"))) %>%
  arrange(Outcome)

# plot
png(file.path(op, "CV3_class_1_weighted_forest_RR.png"), width = 504, height = 387)
p <- ggplot(result_CV3_class_1_s, aes(x = RR, y = Outcome)) +
  geom_point(size = 3) +
  scale_x_log10(limits = c(10**(-0.6), 10), labels = label_number()) +
  scale_y_discrete(limits = rev(levels(factor(result_CV3_class_1_s$Outcome))),
                   labels = rev(c("Any allergen", "Inhalant", "Food", "Milk", "F2",
expression(alpha*"-lactalbumin"), expression(beta*"-lactoglobulin")))) +
  geom_errorbarh(aes(xmin = lowerCI, xmax = upperCI), height = 0.2) +
  geom_vline(xintercept = 1, linetype = "dashed") +
  theme_minimal() +
  theme(plot.title = element_text(hjust = 0.5),
        axis.title.x = element_text(hjust = 0.5, size = 14),
        axis.text.y = element_text(size = 14)) +
  xlab("Risk Ratio") +
  ylab("")

print(p)
dev.off()
```

```
rm(list = ls())

### eof
```

```
# MARTHA trial
# cox models and plots
# 2026-04-28 meweber
# 2026-06-26 mege

library(survey)
library(dplyr)
library(ggplot2)
library(patchwork)

# load data set
load(file.path(fp, "paper_outcomes_analysis_data.rdata"))
rm(per_milk)
rm(PM)

# define survey design
ate  <- svydesign(ids = ~ Record_ID_trans, weights = ~ ipw.ate, dat = MC)

### function for all plots 2026-06-26 mege
cox <- function(time, event, determinant, design, filename, labels = c("comparator",
"intervention")) {
  fo          <- as.formula(paste("Surv(", time,",", event,") ~ ", determinant))
  mo          <- svycoxph(fo, design = design)
  km          <- svykm(fo, design = design)
  # hazards ratio
  est         <- coef(mo)
  cl          <- confint(mo)
  hr          <- sprintf("%.2f", exp(c(est, cl)))
  p           <- round(summary(mo)$coefficients[, "Pr(>|z|)"], 3)
  HR          <- paste0("HR = ", hr[1], " [", hr[2], "-", hr[3], "]")
  # events per group
  e0          <- data.frame(time=km[["0"]]$time, surv=km[["0"]]$surv, group="0")
  e1          <- data.frame(time=km[["1"]]$time, surv=km[["1"]]$surv, group="1")
  curves      <- rbind(e0,e1)
  # censoring
  cens        <- design$variables[, c(time, event, determinant)]
  names(cens) <- c("time", "event", "group")
  cens        <- subset(cens, event==0 & !is.na(time) & !is.na(group))
  cens$group  <- as.character(cens$group)
  c0          <- subset(cens, group=="0")
  c1          <- subset(cens, group=="1")
  idx         <- findInterval(c0$time, e0$time)
  c0$surv     <- e0$surv[pmax(idx, 1)]
  idx_1       <- findInterval(c1$time, e1$time)
  c1$surv     <- e1$surv[pmax(idx_1, 1)]
  censoring   <- rbind(c0, c1)

  x_max     <- max(curves$time, na.rm = TRUE)
  breaks     <- seq(0, floor(max(curves$time, na.rm = TRUE) / 50) * 50, by = 50)

  P <- ggplot(curves, aes(x = time, y = surv, color = group)) +
        geom_step(linewidth = 1) +
        geom_point(data = censoring,
                   aes(x = time, y = surv, color = group),
                   shape = 3, size = 2) +
    coord_cartesian(xlim = c(0, x_max), ylim = c(0, 1)) +
    scale_x_continuous(breaks = breaks, limits = c(0, x_max), expand = c(0, 0)) +
        scale_color_manual(values = c("0" = "#0072B2", "1" = "#CC79A7"),
                           labels = labels) +
        labs(x = "Time until event",
             y = "Probability",
             color = "Group") +
        annotate("text",
                 x = max(curves$time) * 0.7,
                 y = 0.1,
                 label = HR) +
        theme_minimal() +
        theme( panel.grid = element_blank(),
```

```
                  legend.position = c(0.8, 0.4),
                  legend.background = element_rect(fill = "white", color = "black"),
                  axis.title.x = element_text(size = 14),
                  axis.title.y = element_text(size = 14),
                  axis.text.x = element_text(size = 14),
                  axis.text.y = element_text(size = 14))

  dat        <- design$variables[, c(time, determinant)]
  names(dat) <- c("time", "group")
  dat$weight <- weights(design)
  dat$group  <- as.character(dat$group)
  dat        <- subset(dat, !is.na(dat$time))

  risk0      <- sapply(breaks, function(t) {sum(dat$weight[dat$group == "0" & dat$time >= t])})
  risk1      <- sapply(breaks, function(t) {sum(dat$weight[dat$group == "1" & dat$time >= t])})

  risk0      <- round(risk0)
  risk1      <- round(risk1)

  risk       <- data.frame(time = rep(breaks, 2), group = rep(c("0", "1"), each =
length(breaks)), n = c(risk0, risk1))

   risk$group <- factor(risk$group, levels = c("0", "1"), labels = labels)

  risk_plot <- ggplot(risk, aes(x = time, y = group)) +
    geom_text(aes(label = n), size = 4) +
    scale_x_continuous(breaks = breaks, limits = c(0, x_max), expand = c(0, 0)) +
    scale_y_discrete(drop = FALSE, labels = labels) +
    labs(x = NULL, y = NULL) +
    coord_cartesian(ylim = c(0.5, 2.8), clip = "off") +
    theme_minimal() +
    theme(axis.title = element_blank(),
          axis.text.x = element_blank(),
          axis.ticks.x = element_blank(),
          axis.text.y = element_text(size = 12,
                                     margin = margin(r = 12)),
      panel.grid = element_blank(),
      plot.margin = margin(t = 5, r = 5, b = 5, l = 5))

  R <- risk_plot +
    labs(title = "No. at risk") +
    theme(plot.title = element_text(hjust = 0.1, face = "plain", size = 11,
                                    margin = margin(b = -5)),
          plot.title.position = "plot",
          plot.margin = margin(l = 5, r = 5, t = 0, b = 5))

  final_plot <- P / R +
    plot_layout(heights = c(4, 1))

  # create png
  png(file.path(op, paste0(filename, ".png")), width=300, height=300)
  # print(P)
  print(final_plot)
  dev.off()
}

cox(time="time", event="event", determinant="verum", design=ate,
filename="asthma_intervention")

cox(time="time", event="event", determinant="verum", design=subset(ate, parental_asthma == 1),
filename="asthma_intervention_parental_asthma")

cox(time="time_neuroderm", event="event_neuroderm", determinant="verum", design=ate,
filename="eczema_intervention")

rm(list = ls())

### eof
```

```
# MARTHA trial
# plot respiratory infetions
# 2023-05-01 meweber

library(reshape2)

# load data set
load(file.path(fp, "paper_outcomes_analysis_data.rdata"))
rm(per_milk)
rm(PM)

W                               <- subset(MC, select = c(substr(names(MC),1,1) %in% c("R", 
"W")))
M                               <- subset(MC, select = c(substr(names(MC),1,1) != "W"))

inf.vars                        <- NULL
for (v in c("cough", "rhinitis")) {
  inf.vars                      <- c(inf.vars, names(W)[grep(v, names(W))])
}

molten            <- reshape2::melt(W, id.vars = c("Record_ID_trans"), measure.vars = inf.vars, 
na.rm = TRUE)
molten$week       <- as.numeric(substr(molten$variable, 2, 4))
molten$variable   <- as.character(molten$variable)
molten$variable   <- substr(molten$variable, 6, nchar(molten$variable))
molten$value      <- as.numeric(molten$value)
molten$dicho      <- as.numeric(molten$value>0)

molten                          <- merge(molten, M[, c("Record_ID_trans", 
"rounded_birthdate")], all.x=TRUE)

molten$actual_date              <- molten$rounded_birthdate + 7* molten$week -1

weekly_prevalence               <- reshape2::dcast(molten, actual_date ~ variable, 
fun.aggregate = mean, value.var = "dicho")

lockdown_starts <- as.Date(c("2020-03-22", "2020-11-02", "2021-11-24"))
lockdown_ends   <- as.Date(c("2020-06-01", "2021-02-07", "2021-12-15"))

png(file.path(op, paste("Respiratory infections in MARTHA.png")), width = 800, height = 400)
plot(weekly_prevalence$actual_date[weekly_prevalence$actual_date < "2023-06-30"], 
weekly_prevalence$rhinitis[weekly_prevalence$actual_date < "2023-06-30"], type="l", 
ylim=c(0,1), xlab="Year", ylab="Proportion", main="Respiratory infections in MARTHA", cex.axis 
= 1.2, cex.lab = 1.2)
lines(weekly_prevalence$actual_date[weekly_prevalence$actual_date < "2023-06-30"], 
weekly_prevalence$cough[weekly_prevalence$actual_date < "2023-06-30"], type="l", col=2)
for (i in seq_along(lockdown_starts)) {
  rect(xleft = lockdown_starts[i], xright = lockdown_ends[i],
       ybottom = par("usr")[3], ytop = par("usr")[4],
       col = rgb(0.5, 0.5, 0.5, 0.5), border = NA)
}
abline(h = 0.3, lty = 2)
legend("topright", c("rhinitis", "cough"), bg = "white", fill=1:2, cex = 1.2)
dev.off()

rm(list = ls())

### eof
```

```
# MARTHA trial
# rhinitis/cough RR over birth date
# 2026-02-04 meweber, mege

library(dplyr)
library(survey)
library(ggplot2)
library(scales)

load(file.path(fp, "paper_outcomes_analysis_data.rdata"))
rm(per_milk)
rm(PM)

W                               <- subset(MC, select = c(substr(names(MC),1,1) %in% c("R",
"W")))

inf.vars                        <- NULL
for (v in c("cough", "rhinitis", "Rhin0.3")) {
  inf.vars                      <- c(inf.vars, names(W)[grep(v, names(W))])
}

molten              <- reshape2::melt(W, id.vars = c("Record_ID_trans"), measure.vars = inf.vars,
na.rm = TRUE)
molten$week         <- as.numeric(substr(molten$variable, 2, 4))
molten$variable     <- as.character(molten$variable)
molten$variable     <- substr(molten$variable, 6, nchar(molten$variable))
molten$value        <- as.numeric(molten$value)

molten                          <- merge(molten, MC[, c("Record_ID_trans",
"rounded_birthdate")], all.x=TRUE)

molten$actual_date              <- molten$rounded_birthdate + 7* molten$week -1

W                               <- reshape2::dcast(molten, Record_ID_trans + week + actual_date
~ variable, fun.aggregate=mean, fill = as.numeric(NA))

W                               <- merge(W, MC[, c("Record_ID_trans", "ipw.ate", "verum",
"rounded_birthdate")])

prev03  <- svydesign(ids = ~ Record_ID_trans, weights = ~ ipw.ate, dat = subset(W, Rhin0.3==1))

# Rhinitis
{
  MC_sorted <- MC[order(MC$rounded_birthdate, MC$Record_ID_trans),]
  window_size <- 65
  step_size   <- 1  # controls overlap

  starts <- seq(1, nrow(MC_sorted) - window_size + 1, by = step_size)

  result <- do.call(
    rbind,
    lapply(starts, function(i) {

      dat_sub <- MC_sorted[i:(i + window_size - 1), ]
      des_sub <- subset(prev03, Record_ID_trans %in% dat_sub$Record_ID_trans)
      fit     <- svyglm(rhinitis ~ verum, design = des_sub, family = quasipoisson(link="log"))
      est     <- coef(summary(fit))["verum1", ]
      beta    <- est["Estimate"]
      ci      <- confint(fit)

      data.frame(start = min(dat_sub$rounded_birthdate),
                 end   = max(dat_sub$rounded_birthdate),
                 median= median(dat_sub$rounded_birthdate),
                 RR    = exp(beta),
                 lower = exp(ci["verum1", 1]),
                 upper = exp(ci["verum1", 2]),
                 p     = est["Pr(>|t|)"])
    }))
```

```
    png(file.path(op, paste("Rhinitis_per_birthdate.png")), width = 800, height = 400)
    plot(as.Date(result$median),result$RR, ylim=c(0.5, 1.6), type="b", xaxt="n", xlab="Birth date", ylab="RR")
  axis(1, at=c(as.Date("2019-04-01"),as.Date("2019-07-01"),as.Date("2019-10-01"),as.Date("2020-01-01"),as.Date("2020-04-01"),as.Date("2020-07-01"),as.Date("2020-10-01"),as.Date("2021-01-01")),labels = c("2019-04","2019-07","2019-10","2020-01","2020-04","2020-07","2020-10","2021-01"))
  lines(as.Date(result$median),result$upper)
  abline(h=1)
  lines(as.Date(result$median),result$lower)
  dev.off()
  }

# Cough
{
  MC_sorted <- MC[order(MC$rounded_birthdate, MC$Record_ID_trans),]
  window_size <- 65
  step_size   <- 1  # controls overlap

  starts <- seq(1, nrow(MC_sorted) - window_size + 1, by = step_size)

  result <- do.call(
    rbind,
    lapply(starts, function(i) {

      dat_sub <- MC_sorted[i:(i + window_size - 1), ]
      des_sub <- subset(prev03, Record_ID_trans %in% dat_sub$Record_ID_trans)
      fit     <- svyglm(cough ~ verum, design = des_sub, family = quasipoisson(link="log"))
      est     <- coef(summary(fit))["verum1", ]
      beta    <- est["Estimate"]
      ci      <- confint(fit)

      data.frame(start = min(dat_sub$rounded_birthdate),
                 end   = max(dat_sub$rounded_birthdate),
                 median= median(dat_sub$rounded_birthdate),
                 RR    = exp(beta),
                 lower = exp(ci["verum1", 1]),
                 upper = exp(ci["verum1", 2]),
                 p     = est["Pr(>|t|)"])
    }))

  png(file.path(op, paste("Cough_per_birthdate.png")), width = 800, height = 400)
  plot(as.Date(result$median),result$RR, ylim=c(0.5, 1.6), type="b", xaxt="n", xlab="Birth date", ylab="RR")
  axis(1, at=c(as.Date("2019-04-01"),as.Date("2019-07-01"),as.Date("2019-10-01"),as.Date("2020-01-01"),as.Date("2020-04-01"),as.Date("2020-07-01"),as.Date("2020-10-01"),as.Date("2021-01-01")),labels = c("2019-04","2019-07","2019-10","2020-01","2020-04","2020-07","2020-10","2021-01"))
  lines(as.Date(result$median),result$upper)
  abline(h=1)
  lines(as.Date(result$median),result$lower)
  dev.off()
}

rm(list = ls())

### eof
```

```
# MARTHA trial
# analysis of fatty acids in milk and blood
# 2025-07-15 meweber

library(compositions)
library(dplyr)
library(ggplot2)
library(survey)

load(file.path(fp, "paper_outcomes_analysis_data.rdata"))
rm(PM)

omega.list <- c("C18.3n.3", "C20.5n.3", "C22.5n.3", "C18.2n.6", "C20.4n.6", "C22.5n.6",
"C22.6n.3")

# milk
{
  # set order correctly
    per_milk$Sample <- factor(per_milk$Sample, levels = c("G", "T"))

    result_milk <- data.frame(NULL)
    for(A in omega.list) {
      fo <- as.formula(paste(A, "~ Sample"))
      t  <- t.test(fo, per_milk)
      result_milk <- rbind(result_milk, data.frame(A = A, mean_diff = t$estimate[1] -
t$estimate[2] , lowerCI = t$conf.int[1], upperCI = t$conf.int[2], p = format(round(t$p.value,
4), scientific = FALSE)))
    }

    result_milk <- result_milk %>%
      mutate(A = factor(A, levels = c("C18.3n.3", "C20.5n.3", "C22.5n.3", "C22.6n.3",
"C18.2n.6", "C20.4n.6", "C22.5n.6"))) %>%
      arrange(A)

    png(file.path(op, paste("fa_milk_forest.png")), width = 614, height = 412)
    p <- ggplot(result_milk, aes(x = mean_diff, y = A)) +
      geom_point(size = 3) +
      scale_y_discrete(limits = rev(levels(factor(result_milk$A))),
                       labels = rev(c("C18:3-n3", "C20:5-n3", "C22:5-n3", "C22:6-n3", "C18:2-
n6", "C20:4-n6", "C22:5-n6"))) +
      geom_errorbarh(aes(xmin = lowerCI, xmax = upperCI), height = 0.2) +
      geom_vline(xintercept = 0, linetype = "dashed") +
      theme_minimal() +
      theme(plot.title = element_text(hjust = 0.5),
            axis.text.y = element_text(size = 18)) +
      labs(x = "", y = "")
    print(p)
    dev.off()
}

# blood
{
  ate        <- svydesign(ids = ~ LID_trans, weights = ~ ipw.ate, dat = MC)

  result_blood_all <- data.frame(NULL)
  for(A in omega.list) {
    fo <- as.formula(paste(A, "~ verum"))
    t  <- svyttest(fo, design = ate)
    est <- t$estimate
    ci  <- t$conf.int
    result_blood_all <- rbind(result_blood_all, data.frame(A = A, mean_diff = est, lowerCI =
ci[1], upperCI = ci[2], mean_verum = svyby(~get(A), ~verum, design = ate, svymean)[2,2],
mean_placebo = svyby(~get(A), ~verum, design = ate, svymean)[1,2], p = format(round(t$p.value,
4), scientific = FALSE)))
  }

  result_blood_all <- result_blood_all %>%
    mutate(A = factor(A, levels = c("C18.3n.3", "C20.5n.3", "C22.5n.3", "C22.6n.3", "C18.2n.6",
```

```
"C20.4n.6", "C22.5n.6"))) %>%
    arrange(A)

  # forest plot
  {
    png(file.path(op, paste("fa_blood_forest_all_weighted.png")), width = 614, height = 412)
    p <- ggplot(result_blood_all, aes(x = mean_diff, y = A)) +
      geom_point(size = 3) +
      scale_y_discrete(limits = rev(levels(factor(result_blood_all$A))),
                       labels = rev(c("C18:3-n3", "C20:5-n3", "C22:5-n3", "C22:6-n3", "C18:2-
n6", "C20:4-n6", "C22:5-n6"))) +
      geom_errorbarh(aes(xmin = lowerCI, xmax = upperCI), height = 0.2) +
      geom_vline(xintercept = 0, linetype = "dashed") +
      theme_minimal() +
      theme(plot.title = element_text(hjust = 0.5),
            axis.title.x = element_text(hjust = 0.1),
            axis.text.y = element_text(size = 18)) +
      xlab("") +
      ylab("")
    print(p)
    dev.off()
  }

}

rm(list = ls())

### eof
```

```
# Martha trial
# PASTURE scores on MARTHA data
# 2026-05-20 mege, meweber

library(survey)
library(ggplot2)

load(file.path(fp, "paper_outcomes_analysis_data.rdata"))
rm(per_milk)
rm(PM)

ate           <- svydesign(ids = ~ LID_trans, weights = ~ ipw.ate, dat = MC)

# intervention
{
  model <- svyglm(M_diff ~ as.factor(verum), design = subset(ate, parental_asthma == 1))
  MD    <- sprintf("%.2f", coef(summary(model))[2,1])
  lower <- sprintf("%.2f", confint(model)[2,1])
  upper <- sprintf("%.2f", confint(model)[2,2])

  png(file.path(op, paste("bps_verum_fam_hist.png")), width = 557, height = 425)
  svyboxplot(M_diff ~ as.factor(verum), design = subset(ate, parental_asthma == 1), xaxt = "n",
ylim = c(-0.6, 0.8), cex.axis = 1.5 , cex.lab = 1.5)
  axis(1, at = c(1, 2), labels = c("comparator", "intervention"), cex.axis = 1.5)
  text(x = 1.5, y = 0.75, labels = paste0("MD = ", MD, " [", lower, ";", upper, "]"), cex =
1.5)
  dev.off()
}

# asthma
{
  model <- svyglm(M_diff ~ as.factor(event), design = subset(ate, parental_asthma == 1))
  MD    <- sprintf("%.2f", coef(summary(model))[2,1])
  lower <- sprintf("%.2f", confint(model)[2,1])
  upper <- sprintf("%.2f", confint(model)[2,2])

  png(file.path(op, paste("bps_asthma_fam_hist.png")), width = 557, height = 425)
  svyboxplot(M_diff ~ as.factor(event), design = subset(ate, parental_asthma == 1), xlab =
"Asthma diagnosis", xaxt = "n", ylim = c(-0.6, 0.8), cex.axis = 1.5 , cex.lab = 1.5)
  axis(1, at = c(1, 2), labels = c("no", "yes"), cex.axis = 1.5)
  text(x = 1.5, y = 0.75, labels = paste0("MD = ", MD, " [", lower, ";", upper, "]"), cex =
1.5)
  dev.off()
}

rm(list = ls())

### eof
```

```
# MARTHA trial
# Distribution of demographics variable with survey weights
# 2025-06-30 meweber

library(survey)

load(file.path(fp, "paper_outcomes_analysis_data.rdata"))
rm(per_milk)
rm(PM)

# source applicable functions
source(file.path(fp_prg, "functions_svy.r"))

compare.var <- c("female_gender", "C_sectio", "antibiotics_birth_mother_d", "antibiotics_ever",
"any_smoking",
                  "sibs", "daycare", "contact_animals", "parental_asthma", "maternal_asthma",
                 "paternal_asthma", "parental_atopic_eczema",
                 "parental_hay_fever", "parental_atopy", "parental_milk_allergy",
"p_birth_out_G",
                 "school10_f", "school10_m", "rural", "pregnancy_cowsmilk",
                 "any_cowsmilk_before_intervention", "pre_study_formula",
"previous_disease_atopic_eczema_d",
                 "adh_1_year", "additional_milk_d")

# change vector with discrete variables to type factor
MC[, compare.var]         <- lapply(MC[, compare.var], as.factor)

num.vars <- c("full_week_preg", "birth_weight", "age_mother", "breastfeeding_duration",
              "age_CV1", "age_initiation", "age_end_intervention", "intervention_dur_w",
"additional_milk")

name.var                  <- data.frame(V = c(num.vars),
                                        N = c("Completed weeks of pregnancy", "Birth weight
(kg)",
                                              "Maternal age (years)",  "Breastfeeding duration
(weeks)",
                                              "Age at CV1", "Age at start of intervention
(weeks)",
                                              "Age at end of intervention (weeks)",
                                              "Duration of intervention (weeks)",
                                               "Additional milk consumption (weeks)"))

quartile                  <- c("breastfeeding_duration", "additional_milk")

# percentiles
num.vars.per                   <- c("CV1_w", "CV1_h", "CV2_w", "CV2_h", "CV3_w", "CV3_h")

name.var.per                   <- data.frame(V = c(num.vars.per),
                                        N = c("Weight percentiles at CV1",
                                              "Height percentiles at CV1",
                                              "Weight percentiles at CV2",
                                              "Height percentiles at CV2",
                                              "Weight percentiles at CV3",
                                              "Height percentiles at CV3"))

quartile.mean                  <- NULL

## define svy design
design.ate                <- svydesign(ids= ~1, weights= ~ipw.ate, dat = MC)

table.1.ate               <- TA.svy(group="verum", var.list= compare.var, dset = MC, design =
design.ate)

table.1.ate.formated              <- as.data.frame(table.1.ate$Variable)

table.1.ate.formated$complete     <- paste0(table.1.ate$Complete_n, " (",
table.1.ate$Complete_pct, ")")
table.1.ate.formated$intervention <- paste0(table.1.ate$Test.milk_n, " (",
```

```
table.1.ate$Test.milk_pct, ")")
table.1.ate.formated$comparator   <- paste0(table.1.ate$Control.milk_n, " (",
table.1.ate$Control.milk_pct, ")")

table.1a.ate              <- NULL
for(V in name.var$V)
  table.1a.ate           <- rbind(table.1a.ate, tab.num.svy(V, "verum", MC,name.var,
design.ate))

colnames(table.1a.ate)    <- c("Variable", "CC", "Test.milk \n (median and range \n except
breastfeeding_dur, add_milk, num_sibs)", "Control.milk \n (median and range \n except
breastfeeding_dur, add_milk, num_sibs)")
table.1a.ate              <- data.frame(table.1a.ate, stringsAsFactors = FALSE)

table.1a.ate.per              <- NULL
for(V in name.var.per$V)
  table.1a.ate.per           <- rbind(table.1a.ate.per, tab.num.svy.mean(V, "verum",
MC,name.var.per, design.ate))

colnames(table.1a.ate.per)    <- c("Variable", "CC", "Test.milk \n (mean and standard
deviation)", "Control.milk \n (mean and standard deviation)")
table.1a.ate.per              <- data.frame(table.1a.ate.per, stringsAsFactors = FALSE)


write.csv2(table.1.ate.formated, file = file.path(op, paste("table_1_ate.csv")))
write.csv2(table.1a.ate, file = file.path(op, paste("table.1a.ate.csv")))
write.csv2(table.1a.ate.per, file = file.path(op, paste("table.1a.ate.per.csv")))

rm(list = ls())

#### eof
```

```
# MARTHA trial
# prepare SMD plot
# 2024-01-05 meweber

library(survey)
library(tableone)
library(reshape2)
library(ggplot2)
library(useful)

load(file.path(fp, "paper_outcomes_analysis_data.rdata"))
rm(per_milk)
rm(PM)

uw                                <- svydesign(ids = ~ Record_ID_trans, weights = ~ 1, dat = MC)
ate                               <- svydesign(ids = ~ Record_ID_trans, weights = ~ ipw.ate, dat
= MC)

var.list                          <- c("present_weight_gramm", "sibs_hay_fever",
"paternal_atopic_eczema",
                                       "contact_animals",
                                       "any_cowsmilk_before_intervention", "maternal_atopy",
"rural",
                                       "daycare", "any_smoking",
                                       "paternal_asthma", "parental_asthma",
"paternal_hay_fever",
                                       "female_gender", "parental_hay_fever", "parental_atopy",
"age_CV1",
                                       "birth_weight", "full_week_preg", "number_siblings_2_1")

tabUw                      <- svyCreateTableOne(vars = var.list, strata = "verum", data = uw,
test = FALSE)
tabAte                     <- svyCreateTableOne(vars = var.list, strata = "verum", data = ate,
test = FALSE)

dataPlot                   <- data.frame(variable  = c("Weight at CV1 (gr)", "Sibs hay fever",
                                                       "Paternal atopic eczema", "Contact
animals",
                                                       "Cowsmilk before intervention", "Maternal
atopy",
                                                       "Rural living",
                                                       "Daycare", "(Any) smoking", "Paternal
asthma",
                                                       "Parental asthma", "Paternal hay fever",
                                                       "Female gender",  "Parental hay fever",
                                                       "Parental atopy", "Age at CV1",
                                                       "Birth weight", "Completed weeks of
pregnancy",
                                                       "Number siblings"),
                                         Unweighted = as.numeric(ExtractSmd(tabUw)),
                                         IPTW = as.numeric(ExtractSmd(tabAte)))

dataPlotMelt               <- melt(data = dataPlot, id.vars = c("variable"),
                                   variable.name = "Method",
                                   value.name    = "SMD")

varNames                   <- as.character(dataPlot$variable)[order(- dataPlot$Unweighted)]

dataPlotMelt$variable      <- factor(dataPlotMelt$variable, levels = varNames)

png(file.path(op, paste("SMD_plot_flipped.png")), width = 1200, height = 800)
p <- ggplot(data = dataPlotMelt,
       mapping = aes(x = variable, y = SMD, group = Method, color = Method)) +
  geom_line() +
  geom_point(aes(shape = Method, color = Method)) +
  geom_hline(yintercept = 0.1, color = "black", linewidth = 0.1) +
  theme_bw() +
  theme(legend.key = element_blank(),
```

```
        axis.title.x = element_text(size = 14),
        axis.title.y = element_text(size = 14),
        legend.text = element_text(size = 14),
        legend.title = element_text(size = 14),
        axis.text = element_text(size = 12),
        axis.text.x = element_text(angle = 45, hjust = 1, size = 14),
        axis.text.y = element_text(size = 14)) +
ylab("Standardized difference") +
  xlab("Variable")+
  scale_x_discrete(labels = c("Female gender", "Completed weeks \n of pregnancy",
                              "Parental hay fever",
                              "Daycare", "(Any) smoking", "Number siblings", "Parental atopy",
                              "Birth weight", "Paternal asthma", "Parental asthma",
                              "Age at CV1", "Siblings hay fever", "Weight at CV1",
                              "Paternal atopic eczema", "Paternal hay fever",
                              "Contact animals", "Cowsmilk before intervention",
                              "Maternal atopy", "Rural living"))
print(p)
dev.off()

rm(list = ls())

### eof
```

```
# MARTHA trial
# plot milk consumption in children without parental asthma, with parental asthma
# 2026-03-24 meweber

load(file.path(fp, "paper_outcomes_analysis_data.rdata"))
rm(per_milk)
rm(PM)

# subset meta.weekly.trans in weekly data
W                 <- subset(MC, select = c(substr(names(MC),1,1) %in% c("R", "W")))

studymilk.vars    <- names(W)[grep("studymilk_quant", names(W))]

molten          <- reshape2::melt(W, id.vars = c("Record_ID_trans"), measure.vars =
studymilk.vars, nar.rm = TRUE)
molten$week     <- as.numeric(substr(molten$variable, 2, 4))
molten$variable <- as.character(molten$variable)
molten$var.new  <- substr(molten$variable, 6, nchar(molten$variable))
W               <- reshape2::dcast(subset(molten, !is.na(value)), Record_ID_trans + week ~
var.new, fill = 0)

milkquant       <- reshape2::melt(W, id.vars= c("Record_ID_trans", "week") ,
measure.vars="studymilk_quant", na.rm=TRUE, value.name = "studymilk_quant")

milkquant       <- merge(milkquant, MC[, c("Record_ID_trans", "parental_asthma", "start_weeks",
"end_weeks")], all.x = TRUE)

milkquant$entire_bottle <- ifelse(milkquant$studymilk_quant >= 2, 1, 0)
milkquant$half_bottle   <- ifelse(milkquant$studymilk_quant >= 1, 1, 0)

# filter for applicable weeks
milkquant        <- subset(milkquant, week >= start_weeks & week <= end_weeks)

milkquant$start_weeks  <- NULL
milkquant$end_weeks    <- NULL
milkquant$variable     <- NULL

png(file.path(op, paste("week_milk_consump_stratified.png")), width = 548, height = 430)
y <- prop.table(table(milkquant$week[milkquant$parental_asthma == 0],
milkquant$entire_bottle[milkquant$parental_asthma == 0]), 1)[,2]
x <- as.numeric(names(y))
plot(x,y, type="l", xlab = "week of life", ylab = "proportion with entire bottle", xlim = c(25,
160), ylim = c(0, 1), cex.lab=1.3)

Y <- prop.table(table(milkquant$week[milkquant$parental_asthma == 1],
milkquant$entire_bottle[milkquant$parental_asthma == 1]), 1)[,2]
X <- as.numeric(names(Y))
lines(X,Y, col = 2)

abline(v = c(52, 78, 104, 130, 156), col='grey')
legend("top", c("without parental asthma", "with parental asthma"), fill=1:2, box.lty=0, bg =
"white")

dev.off()

rm(list = ls())

### eof
```

```
# MARTHA trial
# rhinitis incidence plot per age half
# 2026-05-01 meweber

load(file.path(fp, "paper_outcomes_analysis_data.rdata"))
rm(per_milk)
rm(PM)

W                                <- subset(MC, select = c(substr(names(MC),1,1) %in% c("R",
"W")))
M                                <- subset(MC, select = c(substr(names(MC),1,1) != "W"))

inf.vars                         <- NULL
for (v in c("rhinitis")) {
  inf.vars                       <- c(inf.vars, names(W)[grep(v, names(W))])
}

molten             <- reshape2::melt(W, id.vars = c("Record_ID_trans"), measure.vars = inf.vars,
na.rm = TRUE)
molten$week        <- as.numeric(substr(molten$variable, 2, 4))
molten$variable    <- as.character(molten$variable)
molten$variable    <- substr(molten$variable, 6, nchar(molten$variable))
molten$value       <- as.numeric(molten$value)
molten$dicho       <- as.numeric(molten$value>0)

molten                           <- merge(molten, M[, c("Record_ID_trans", "rounded_birthdate",
"young_kids")], all.x=TRUE)

plot_half <- function(H) {
  WP <- reshape2::dcast(subset(molten,young_kids == H), week ~ variable, fun.aggregate = mean,
value.var = "dicho")
  WP_sorted <- WP[order(WP$week), ]
  window_size <- 10
  step_size   <- 1  # controls overlap
  starts <- seq(1, nrow(WP_sorted) - window_size + 1, by = step_size)
  result <- do.call(
    rbind,
    lapply(starts, function(i) {

      dat_sub <- WP_sorted[i:(i + window_size - 1), ]

      data.frame(start = min(dat_sub$week),
                 end   = max(dat_sub$week),
                 median= median(dat_sub$week),
                 prev  = mean(dat_sub$rhinitis))
    }))
  plot(WP$week, WP$rhinitis, type="l",  ylim=c(0,.6), xlab="Age in weeks", ylab="Prevalence",
main=ifelse(H==1, "Younger half", "Older half"), cex.axis = 1.5, cex.lab = 1.5, cex.main=1.5)
  lines(result$median,result$prev, col=2, lwd=2)
  abline(h=.3,lty=2)
}

png(file.path(op, paste("rhinitis in older halfs.png")), width = 800, height = 400)
plot_half(0)
dev.off()

png(file.path(op, paste("rhinitis in younger halfs.png")), width = 800, height = 400)
plot_half(1)
dev.off()

rm(list = ls())

### eof
```

```
# MARTHA trial
# table IgE CV3 class 1 and 2
# 2026-07-16 meweber

library(survey)
library(dplyr)

# load data set
load(file.path(fp, "paper_outcomes_analysis_data.rdata"))
rm(per_milk)
rm(PM)

IgE.list   <- names(MC)[grep("cl2", names(MC))]
IgE.list.1 <- names(MC)[grep("cl1", names(MC))]

# define survey designs
ate                <- svydesign(ids = ~ LID_trans, weights = ~ ipw.IgE_sub, dat = subset(MC,
!is.na(ipw.IgE_sub)))

result_CV3_class_2 <- data.frame(NULL)
for(A in IgE.list) {
  fo                 <- as.formula(paste(A, "~ as.numeric(verum)"))
  model              <- svyglm(fo, design = ate, family = quasipoisson(link = "log"))
  RR                 <- sprintf("%.2f", exp(coef(model)[2]))
  CI                 <- exp(confint(model)[2,])
  fl                 <- as.formula(paste0("~", A, "+verum"))
  X                  <- svytable(fl, ate)
  result_CV3_class_2 <- rbind(result_CV3_class_2, data.frame(A, paste0(round(X["1","1"]), " (",
round(100* (round(X["1","1"])/sum(round(X[,"1"])))), "%)"), paste0(round(X["1","0"]), " (",
round(100* (round(X["1","0"])/sum(round(X[,"0"])))), "%)"), summary(model)$coef[2,
c("Estimate")], summary(model)$coef[2, c("Pr(>|t|)")], RR, CI[1], CI[2], paste0(RR, " ", "[",
sprintf("%.2f", CI[1]), " - ", sprintf("%.2f", CI[2]), "]")))
}

names(result_CV3_class_2)          <- c("Outcome", "intervention", "comparator", "Estimate",
"p", "RR", "lowerCI", "upperCI", "RR_CI")

result_CV3_class_1 <- data.frame(NULL)
for(A in IgE.list.1) {
  fo                 <- as.formula(paste(A, "~ as.numeric(verum)"))
  model              <- svyglm(fo, design = ate, family = quasipoisson(link = "log"))
  RR                 <- sprintf("%.2f", exp(coef(model)[2]))
  CI                 <- exp(confint(model)[2,])
  fl                 <- as.formula(paste0("~", A, "+verum"))
  X                  <- svytable(fl, ate)
  result_CV3_class_1 <- rbind(result_CV3_class_1, data.frame(A, paste0(round(X["1","1"]), " (",
round(100* (round(X["1","1"])/sum(round(X[,"1"])))), "%)"), paste0(round(X["1","0"]), " (",
round(100* (round(X["1","0"])/sum(round(X[,"0"])))), "%)"), summary(model)$coef[2,
c("Estimate")], summary(model)$coef[2, c("Pr(>|t|)")], RR, CI[1], CI[2], paste0(RR, " ", "[",
sprintf("%.2f", CI[1]), " - ", sprintf("%.2f", CI[2]), "]")))
}

names(result_CV3_class_1)          <- c("Outcome", "intervention", "comparator", "Estimate",
"p", "RR", "lowerCI", "upperCI", "RR_CI")

result_CV3_class_2$RR <- as.numeric(result_CV3_class_2$RR)
result_CV3_class_1$RR <- as.numeric(result_CV3_class_1$RR)

rownames(result_CV3_class_2) <- NULL
rownames(result_CV3_class_1) <- NULL

write.csv2(result_CV3_class_2, file = file.path(op, paste("IgE_CV3_class_2.csv")))
write.csv2(result_CV3_class_1, file = file.path(op, paste("IgE_CV3_class_1.csv")))

rm(list = ls())

### eof
```

```
# MARTHA trial
# overview asthma cases
# 2026-05-07 meweber

library(dplyr)
library(survey)

# load data set
load(file.path(fp, "paper_outcomes_analysis_data.rdata"))
rm(per_milk)
rm(PM)


# asthma
{
  AC       <- subset(MC, MC$event == 1, select = c("trigger", "verum", "female_gender",
"parental_asthma"))

  tab      <- as.data.frame(table("Type_of_event" = AC$trigger, "Study_arm" = AC$verum, "Sex" =
AC$female_gender, "Parental_asthma" = AC$parental_asthma))

  names(tab)[5] <- "Count"

  tab <- tab[order(tab$Type_of_event, tab$Study_arm, tab$Sex, tab$Parental_asthma), ]

  tab <- tab %>%
    mutate(Type_of_event = replace(as.character(Type_of_event), Type_of_event == "asthma",
"Asthma diagnosis"))
  tab <- tab %>%
    mutate(Type_of_event = replace(as.character(Type_of_event), Type_of_event == "bronchitis",
"Bronchitis twice"))

  tab <- tab %>%
    mutate(Study_arm = replace(as.character(Study_arm), Study_arm == 0, "Comparator"))
  tab <- tab %>%
    mutate(Study_arm = replace(as.character(Study_arm), Study_arm == 1, "Intervention"))

  tab <- tab %>%
    mutate(Sex = replace(as.character(Sex), Sex == 1, "Female"))
  tab <- tab %>%
    mutate(Sex = replace(as.character(Sex), Sex == 0, "Male"))

  tab <- tab %>%
    mutate(Parental_asthma = replace(as.character(Parental_asthma), Parental_asthma == 1,
"Yes"))
  tab <- tab %>%
    mutate(Parental_asthma = replace(as.character(Parental_asthma), Parental_asthma == 0,
"No"))

  write.csv2(tab, file = file.path(op, paste("asthma_cases.csv")))

  rm(AC)
  rm(tab)
}

rm(list = ls())

### eof
```

```
# MARTHA trial
# prepare table for risk ratios with dichotomized birth date
# 2026-02-04 meweber

library(survey)

# load data set
load(file.path(fp, "paper_outcomes_analysis_data.rdata"))
rm(per_milk)
rm(PM)

W                                <- subset(MC, select = c(substr(names(MC),1,1) %in% c("R",
"W")))
M                                <- subset(MC, select = c(substr(names(MC),1,1) != "W"))

inf.vars                         <- NULL
for (v in c("cough", "rhinitis", "Rhin0.3")) {
  inf.vars                       <- c(inf.vars, names(W)[grep(v, names(W))])
}

molten            <- reshape2::melt(W, id.vars = c("Record_ID_trans"), measure.vars = inf.vars,
na.rm = TRUE)
molten$week       <- as.numeric(substr(molten$variable, 2, 4))
molten$variable   <- as.character(molten$variable)
molten$variable   <- substr(molten$variable, 6, nchar(molten$variable))
molten$value      <- as.numeric(molten$value)

molten <- merge(molten, MC[, c("Record_ID_trans", "rounded_birthdate")])
molten$actual_date  <- molten$rounded_birthdate + 7* molten$week -1

MD                 <- reshape2::dcast(molten, Record_ID_trans + week + actual_date ~ variable,
fun.aggregate=mean, fill = as.numeric(NA))

MD                <- merge(MD, MC[, c("Record_ID_trans", "rounded_birthdate", "ipw.ate",
"young_kids", "verum")], all.x=TRUE)

prev03        <- svydesign(ids = ~ Record_ID_trans, weights = ~ ipw.ate, dat = subset(MD,
Rhin0.3==1))

# interaction
estimates <- function(model) {
  RR <- paste0(round(exp(coef(model)), 2)," [", round(exp(confint(model)[,1]),3),"-
",round(exp(confint(model)[,2]),3),"]")
  re <- cbind(round(summary(model)$coef,3), RR)
  re <- re[-1,]
return(re)}

rhin_all    <- estimates(svyglm(as.formula(rhinitis ~ verum*young_kids), design = prev03,
family = quasipoisson(link = "log")))
rhin_young  <- estimates(svyglm(as.formula(rhinitis ~ verum), design = subset(prev03,
young_kids==1), family = quasipoisson(link = "log")))
rhin_old    <- estimates(svyglm(as.formula(rhinitis ~ verum), design = subset(prev03,
young_kids==0), family = quasipoisson(link = "log")))

cough_all   <- estimates(svyglm(as.formula(cough ~ verum*young_kids), design = prev03, family =
quasipoisson(link = "log")))
cough_young <- estimates(svyglm(as.formula(cough ~ verum), design = subset(prev03,
young_kids==1), family = quasipoisson(link = "log")))
cough_old   <- estimates(svyglm(as.formula(cough ~ verum), design = subset(prev03,
young_kids==0), family = quasipoisson(link = "log")))

result <- data.frame(Outcome = "Rhinitis", Subset = "all", Determinant = "Intervention", 'Risk
ratio' = paste(rhin_all[1, 5]), p = paste(rhin_all[1, 4]))

result <- rbind(result, c("Rhinitis", "all", "Younger than median", paste(rhin_all[2, 5]),
paste(rhin_all[2, 4])))
result <- rbind(result, c("Rhinitis", "all", "Interaction term", paste(rhin_all[3, 5]),
paste(rhin_all[3, 4])))
```

```
result <- rbind(result, c("Rhinitis", "Younger than median", "Intervention",
paste(rhin_young[5]), paste(rhin_young[4])))
result <- rbind(result, c("Rhinitis", "Older than median", "Intervention", paste(rhin_old[5]),
paste(rhin_old[4])))

result <- rbind(result, c("Cough", "all", "Intervention", paste(cough_all[1, 5]),
paste(cough_all[1, 4])))
result <- rbind(result, c("Cough", "all", "Younger than median", paste(cough_all[2, 5]),
paste(cough_all[2, 4])))
result <- rbind(result, c("Cough", "all", "Interaction term", paste(cough_all[3, 5]),
paste(cough_all[3, 4])))
result <- rbind(result, c("Cough", "Younger than median", "Intervention",
paste(cough_young[5]), paste(cough_young[4])))
result <- rbind(result, c("Cough", "Older than median", "Intervention", paste(cough_old[5]),
paste(cough_old[4])))

write.csv2(result, file = file.path(op, paste("infections.age.half.prev03.csv")))

rm(list = ls())

#### eof
```

```
# MARTHA trial
# Table on proteins
# 2026-07-23 mege, meweber

# load data set
load(file.path(fp, "paper_outcomes_analysis_data.rdata"))
rm(MC)
rm(per_milk)

PM$intervention_to_raw_pct        <- round(PM$Intervention/PM$raw * 100)
PM$comparator_to_raw_pct          <- round(PM$Comparator/PM$raw * 100)
PM$comparator_to_intervention_pct <- round(PM$Comparator/PM$Intervention * 100)
PM$comparator_to_uht_pct          <- round(PM$Comparator/PM$uht * 100)
PM$intervention_to_comparator     <- round(PM$Intervention/PM$Comparator,2)
PM$raw_to_intervention            <- round(PM$raw/PM$Intervention,2)

PM <- PM[order(PM$comparator_to_intervention_pct),]
PM <- subset(PM, Protein!="Prostaglandin-H2 D-isomerase",
select=c("Protein","intervention_to_raw_pct", "comparator_to_raw_pct",
"comparator_to_intervention_pct"))

write.csv2(PM, file = file.path(op, paste("Protein_table.csv")))

rm(list = ls())

### eof
```
